# Repurposing Shotgun Metagenomic Human Host Reads for Mitochondrial Heteroplasmy Detection

**DOI:** 10.64898/2026.10.05.26364752

**Authors:** Siddharth Singh Tomar, Krishna Khairnar

**Author notes:** Corresponding author: Correspondence to Krishna Khairnar.

## Abstract

**Background:** Shotgun metagenomic pipelines routinely discard host-derived reads, which can exceed 90% of reads in low-biomass clinical samples. Whether these reads support mitochondrial heteroplasmy detection, and at what allele fraction, has not been established.

**Methods:** Shotgun metagenomic data from 47 upper respiratory tract swabs from healthy adults in Nagpur, India, were analysed. Human Reads were recruited against the rCRS and realigned competitively to GRCh38 to remove nuclear mitochondrial segments (NUMTs). Kraken2 was used as a microbial control. Heteroplasmy was called against a pooled, substitution-specific beta-binomial error model and a leave-one-out panel of normals. The detection limit was evaluated from the substitution spectrum against gnomAD v3.1.

**Results:** Complete mitochondrial genomes were recovered from all samples (mean depth 433×, range 9–3,029×). Competitive realignment relocated 3.22% of recruited reads to nuclear loci; microbial cross-mapping removed 0.045%. Error rates varied 12.8-fold across substitution classes, with T>A being the highest. The 134 calls had a Ti/Tv of 0.43, compared with 3.51 in gnomAD, and T>A accounted for 42.5% of calls. T>A allele fractions never exceeded 4.72%, whereas transitions reached 90.81%. At a 5% threshold, T>A calls disappeared, and Ti/Tv rose to 2.20 before plateauing; 16 calls exceeded this limit. Macrohaplogroups were assigned in 46 of 47 samples (M 22, R 23, N 1).

**Conclusions:** Metagenomic host reads support complete mitochondrial recovery, haplogroup typing, and heteroplasmy detection at an allele fraction≥5%. Below this, a T>A-dominated error process persists after NUMT removal that background-fitted models do not capture.

## Introduction

Shotgun metagenomics, originally developed to characterise complex microbial communities in environmental samples, offers a broader and more unbiased approach than 16S amplicon sequencing, as it sequences all DNA present in a sample without amplicon bias. This untargeted strategy is particularly valuable for exploratory compositional profiling and has led to its adoption in clinical research (Quince et al., 2017), where it is used for culture-independent pathogen detection, microbiome characterisation, antimicrobial resistance profiling, and host-response analysis (Chiu & Miller, 2019).

For clinical applications, metagenomic sequencing is typically performed on samples of human origin, including tissues, blood, tumour biopsies, and body fluids or excreta such as urine, stool, endotracheal aspirates, respiratory swabs, and cerebrospinal fluid. As a result, shotgun metagenomic datasets from clinical specimens may contain not only microbial sequences but also variable proportions of host-derived DNA, with host-derived reads contributing >90% of the reads in low-biomass samples such as respiratory swabs and CSF(Chiu & Miller, 2019; Pereira-Marques et al., 2019).

These host-derived DNA sequences, hereafter referred to as host reads, are generally considered uninformative for metagenomic studies focused on microbial profiling. As a result, a large fraction of host reads is filtered or discarded during post-sequencing data analysis (Kalantar et al., 2020). While this filtering is important for accurate microbial composition analysis, it also leads to the loss of potentially valuable host genomic information.

Host reads represent a potentially underutilised source of genomic information, particularly in low-biomass samples such as respiratory swabs (Kim et al., 2024). It is, however, difficult to reconstruct a complete/nearly complete host genome from shotgun reads for genome-wide analysis, but the host reads may contain considerable copies of mitochondrial DNA, owing to its comparatively stable circular structure and high copy number per cell (Sarhan et al., 2025). Mitochondrial DNA is also a practical target for a second reason: its size. At 16,569 bp, the mitochondrial genome can be covered to a reasonable depth by a read fraction that would be far too small for nuclear variant calling (Picardi & Pesole, 2012). The information it carries is correspondingly well characterised. Homoplasmic variants define maternal lineages and explain the human population history (van Oven & Kayser, 2009), while heteroplasmic variants, in which mutant and wild-type DNA coexist within the cell, accumulate with age and have been implicated in mitochondrial disease, cancer and degenerative processes (Gupta et al., 2023; Stewart & Chinnery, 2015; Yuan et al., 2020). This raises the possibility of repurposing host-derived reads from existing shotgun metagenomic datasets for mitochondrial genome analysis and for detecting mitochondrial sequence variation, including heteroplasmic variants.

Repurposing discarded host reads addresses a critical methodological limitation in microbiome research by providing a co-localised, multi-omics data layer. A fundamental bottleneck in tumour microbiome studies is the inability to directly link microbial alterations to host genomic instability (Burns et al., 2018; Okuda et al., 2021). By extracting host heteroplasmy data from the same sample, we attempt to bridge this gap, moving beyond simple correlational observations to generate testable mechanistic hypotheses about the tumour microenvironment. Beyond tumour microbiome profiling, these repurposed host reads provide a foundation for population-level genomic classification based on haplogroup assignments. This allows researchers to correlate microbial signatures with host ancestry and genetic background using data generated from a single sequencing run.

Recovering mitochondrial reads from metagenomic data comes with its own challenges. A major challenge is the interference from nuclear mitochondrial segments (NUMTs). NUMTs frequently align to the mitochondrial reference and appear as low-frequency variants indistinguishable from real heteroplasmy (Laricchia et al., 2022; Wei et al., 2022). NUMT artifacts are pervasive enough that the Genome Aggregation Database (gnomAD™) reports heteroplasmy only above 10% (Laricchia et al., 2022). Another challenge is the inconsistent output of host reads in metagenomic sequencing datasets; host fractions vary by orders of magnitude between samples, so the lowest detectable heteroplasmy level also varies (González et al., 2020). At low allele fractions, heteroplasmy and sequencing error yield the same observation, and distinguishing them requires an error model that accounts for substitution-specific rate differences (Gerstung et al., 2012; Li et al., 2010).

Despite the analytical difficulties discussed above, recovery of host reads from metagenomic sequencing data has several precedents. Off-target reads from whole-exome sequencing have been used to assemble mitochondrial genomes, yielding near-complete sequences from 76% of 1,242 exomes in the 1000 Genomes Project (Diroma et al., 2014), and comparison with Sanger sequencing showed that heteroplasmy levels can be estimated from off-target reads, with error rates comparable to Sanger above 20-fold depth (Griffin et al., 2014). For metagenomes specifically, Tomofuji et al. 2023, showed that host reads in gut metagenomic data carry substantial personal genetic information, sufficient to infer genetic sex and to re-identify individuals against genotype databases (Tomofuji et al., 2023).

The conditions under which heteroplasmy can be inferred from metagenomic host reads have not been established. Published heteroplasmy baselines derive overwhelmingly from European cohorts. (Gupta et al., 2023; Laricchia et al., 2022; Payne et al., 2013), leaving the mitochondrial lineages that predominate in South Asian populations (Chandrasekar et al., 2009), and non-blood tissues are poorly characterised (Samuels et al., 2013). Host reads are increasingly removed before public deposit, using tools such as NCBI’s Human Read Removal Tool, which is designed to be applied as the final step before submission (Katz et al., 2021) archived metagenomes processed in this way cannot be reanalysed for host variation. This practice exists for the right reason: as host reads carry identifying maternal-lineage information, and their reuse raises questions of privacy and consent. (Tomofuji et al., 2023) Retaining this subset under appropriate access controls would cost little and would make host mitochondrial analysis available for studies already underway.

In this proof-of-concept study, we evaluated whether mitochondrial heteroplasmy can be reliably recovered from the host fraction of shotgun metagenomic data, and under what conditions. We analysed 47 nasopharyngeal and oropharyngeal swabs from adults in Nagpur, India, which exhibited a wide range of host DNA content. These samples underwent shotgun metagenomic sequencing on the Illumina platform, and the resulting data were processed through a dedicated bioinformatic pipeline. We characterised mitochondrial DNA recovery as a function of host content, quantified NUMT contamination and microbial cross-mapping, developed an error model tailored to variable sequencing depths, and determined the allele fraction threshold at which heteroplasmic calls can be distinguished from sequencing error.

## 2. Methods

### 2.1. Study cohort and sequencing data

Nasopharyngeal swabs were collected from 48 healthy adult volunteers without known cancer in Nagpur, Maharashtra, India, March-April 2023, as the control arm of a larger upper respiratory tract (URT) shotgun metagenomics study. One sample was excluded due to a pre-sequencing quality-control failure, leaving 47 samples for analysis. Participants in this subset were predominantly male (36/47, 77%) and young (median age 28 years, IQR 25-40, range 22-54), and most resided in urban areas (44/47, 94%). Samples from the control arm of the URT shotgun study were selected because these participants provided informed consent for both genomic analysis and microbiome sequencing during recruitment.

DNA extraction was performed with the QIAamp DNA Microbiome Kit (Catalogue No. 51704), followed by library preparation using the QIAseq FX kit. Samples were sequenced on an Illumina NextSeq 550 system using a 2 × 150 bp high-output kit. While standard metagenomic profiling was conducted via the CZ ID platform (Kalantar et al., 2020). The raw FASTQ files acquired prior to host-read removal were isolated and used as the primary input for all subsequent host genomic analyses

### 2.2. Workflow implementation

The analysis was implemented as a Snakemake (v8.14.0) workflow executed in a Docker container with all software versions pinned. Principal tools were fastp v0.23.4, BWA-MEM v0.7.18, SAMtools v1.20, Kraken2 v2.1.3, Python v3.11.9, pysam v0.22.1, NumPy v1.26.4 and SciPy v1.13.1. The revised Cambridge Reference Sequence (rCRS; NC_012920.1, 16,569 bp) was embedded in the container, and its length was verified at build time. The human reference was GRCh38 (UCSC hg38, without ALT contigs). All parameters were recorded in a single configuration file, which is deposited with the results. https://doi.org/10.5281/zenodo.23034211.

### 2.3. Read preprocessing

Reads were trimmed with fastp using automatic paired-end adapter detection, trailing-end quality trimming (mean quality ≥20), a base-quality threshold of 20, removal of reads with more than 40% low-quality bases, and a minimum length of 50 bp. Trimming was deliberately permissive, to retain the read-end information used by downstream artifact filters.

Mitochondrial reads Recovery used two alignment passes, the first optimised for sensitivity and the second for specificity.

In the first pass, trimmed reads were aligned to the mitochondrial reference (rCRS) alone using BWA-MEM. Properly paired reads mapping to the mitochondrial genome with a mapping quality (MAPQ) ≥30 were retained as candidates. Aligning to the mitochondrial reference alone recruits every read with mitochondrial similarity, including those originating from nuclear mitochondrial segments (NUMTs).

Candidate alignments were name-sorted and converted to paired FASTQ, with singletons discarded. Read-pair counts were verified to be identical in both files, because extracting pairs from a coordinate-sorted alignment desynchronises (R1-R2) mates, and BWA-MEM then produces empty output without reporting an error.

In the second pass, candidate reads were realigned with BWA-MEM to the complete GRCh38 genome, which includes the mitochondrial contig. Competitive alignment lets NUMT-derived reads map to their nuclear locus of origin. Reads mapping to chrM with Mapping Quality (MAPQ) ≥30 were retained. NUMT relocation was quantified per sample from primary alignments only, as the proportion of candidate reads assigned to nuclear chromosomes, together with the principal nuclear destination.

### 2.4. Microbial classification

To test for microbial reads cross-mapping to the mitochondrial genome, reads retained after the second pass were classified with Kraken2 v2.1.3 (Wood et al., 2019) using the standard 8 GB database (Standard-8, build date 2024-05-26) in paired-end mode. Reads assigned to any taxon within Bacteria, Archaea or Viruses were removed; reads classified as human or left unclassified were retained. The default behaviour of Kraken is to discard unclassified and Human reads. Therefore, it discards the target data rather than the microbial reads. Applied to a validation sample, it removed 39,409 of 42,857 reads (92%), with Homo sapiens as the most abundant taxon among the discarded reads. Filtering was based on taxonomy rather than unclassified status because the database contains the human genome, and genuine mitochondrial reads are therefore classified as *Homo sapiens*. We report this explicitly because the failure is silent as the pipeline completes, the output is well-formed, and the loss appears only as an unexplained fall in depth. The workflow was corrected to handle this and now looks up each read’s assigned organism. Discard it only if the organism falls under Bacteria, Archaea, or Viruses; it retains human and unclassified reads.

### 2.5. Coverage and callability

Per-position depth was computed using SAMtools (MAPQ ≥30, all positions reported), yielding mean depth and the fraction of the genome covered at ≥50×, ≥100×, and ≥500×. Because mean depth does not indicate which positions support variant detection, we also computed a callable fraction for each sample. With a minimum of four alternate reads required, detecting a variant at allele fraction *f* requires depth ≥4/*f*. (Cibulskis et al., 2013) The callable fraction at *f* was defined as the proportion of positions meeting this depth, and was reported at *f* = 2%, 3%, 5% and 10%.

### 2.6. Pooled substitution-specific error model

Allele counts were obtained by pileup (base quality ≥20, MAPQ ≥30, deletions excluded). Background error was modelled from positions unlikely to carry true variation: for each position with depth ≥50, each non-reference allele with allele fraction below 0.5% contributed one observation (alternate count, depth). Every non-reference allele was counted separately, not only the most frequent, to avoid preferentially sampling the substitution with the highest error rate at each site.

Observations were pooled across all samples and grouped by substitution type, giving 12 classes (possible combinations of substitution and transversions). Each class was fitted with a beta-binomial distribution using maximum-likelihood estimation (Nelder-Mead optimisation). Classes with fewer than 500 background observations (particularly rare sequencing errors) inherited a global fit across all classes. The model was pooled across samples because per-sample fits collapsed at low depth, where most background positions carry no alternate reads. It was fitted per substitution type, because error rates differed substantially between classes. The concentration parameter (α + β) was bounded at 10 because unbounded optimisation drove it toward values where the beta-binomial tail probability evaluated to exactly zero at every position, silently disabling the significance test.

### 2.7. Site-specific background (panel of normals)

Some positions have local error rates above the substitution-level rate. Because the background was defined as allele fraction below 0.5%, local error cannot be captured by the substitution model. We therefore constructed a panel of normals from the samples itself. For each position and alternate allele, alternate and total read counts were summed across all samples with depth ≥50 at that position. When calling in sample *i*, a site-specific background rate was calculated with that sample left out:

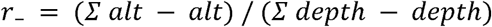

This estimate was used only when at least three other samples covered the position and *r*₋ exceeded the substitution-level rate. A private heteroplasmy contributes nothing to the other samples, so its site rate remains low which is not the case with sequencing error.

### 2.8. Variant calling

At each position with depth ≥50, the most frequent non-reference allele was tested. Two poly-C regions (m.302-315 and m.16184-16193) were excluded because they contain unstable stretches of poly C located in the mitochondrial D-loop. A candidate variant was required to have at least four alternate reads and an allele fraction of at least 1%.

Significance was assessed as the upper-tail probability of the observed alternate count under the substitution-specific beta-binomial model. Where a site-specific background applied, a binomial tail probability at the leave-one-out (r₋ ) rate was also calculated, and the larger of the two probabilities was carried forward. Candidate variants with P < 0.05 passed the significance filter. Strand bias was tested with Fisher’s exact test on forward and reverse-strand counts of the alternate and reference alleles, and candidates with P ≤ 0.01 were rejected.

Variants with an allele fraction of at least 95% were classified as homoplasmic, and those below it as heteroplasmic. The significance and strand-bias filters were applied only to the heteroplasmic class. Each call was assigned to a genomic region: control region (m.16024-576), MT-RNR1 (m.648-1601), MT-RNR2 (m.1671-3229), one of the thirteen protein-coding genes by its annotated coding coordinates, or tRNA/intergenic. Positions between m.3307 and m.15887 that lie outside a coding sequence the seventeen interspersed tRNA genes and the short intergenic segments, together 1,240 bp or 9.9% of that interval were assigned to tRNA/intergenic rather than to protein-coding.

To quantify what competitive realignment contributes, the workflow was re-run for all 47 samples with Pass 2a omitted. Trimming and recruitment were unchanged; because the recruitment reference is the rCRS alone, the Pass 1 output served directly as input to calling. The error model and panel of normals were refitted on the unfiltered alignments, so the comparison is between complete pipelines rather than between read sets. Pass 2b was not carried forward during the final analysis, having affected only 0.045% of reads in the main analysis. **(Table 1)** The details of passes 2a and 2b and their corresponding effects on calls are provided as **Supplementary Table S13**

**Table 1:** Recruitment: aligning against the rCRS alone recruits every read with mitochondrial similarity, including nuclear and microbial sequence. Pass 2a: Competitive alignment against the complete human genome allows NUMT-derived reads to return to their locus of origin. Pass 2b: Tests for microbial cross-mapping and is reported as a verifiable negative control rather than a necessary filter.

| Pass | Reference | Criterion | Reads removed | Effect on calls |
| --- | --- | --- | --- | --- |
| 1. Recruitment | rCRS alone<br>(NC_012920.1, 16,569 bp) | Properly paired, MAPQ ≥30 | — | 3,908,080 reads recruited cohort-wide; NUMT-derived and microbial reads retained by design |
| 2a. Human verification | GRCh38, complete (no ALT contigs) | Aligning to chrM at MAPQ ≥30 | 115,302 reads relocated to nuclear loci (2.95% pooled; per-sample mean 3.22%, range 1.53-6.35%); chr1 predominant in 47/47 | 1,081 → 134 heteroplasmic calls (87.6% removed); 79 → 16 calls above 5% |
| 2b. Microbial classification | Kraken2 Standard-8 (2024-05-26) | Retain human and unclassified; remove Bacteria, Archaea, Viruses | 812 of 1,804,205 read pairs (0.045%); maximum 0.644% in any sample | Negative control; no material effect |

### 2.9. Detection limit

Substitution classes were taken from the per-call substitution field. The observed spectrum of heteroplasmic calls was compared with the variant profile generated by gnomAD v3.1 chrM variants by χ² test against gnomAD proportions across the twelve substitution classes. The transition-to-transversion (Ti/Tv) ratio was computed for all calls and recomputed with a 5% minimum allele-fraction threshold. However, our final statistical detection limit follows the previously described alternate read logic. For example; with four alternate reads required, detection at allele fraction f requires depth ≥ 4/f. At the cohort median depth of 214×, this yielded a detection threshold of around 1.9%, below which the pipeline could not call heteroplasmy. For a better reproducibility across diverse cohorts the detection limit of 5% was selected based on the Ti/Tv ratio optimisation **(Supplementary Table S6)**, Ti/Tv plateaus at 5%. It climbs from 0.43 to 2.20 as the threshold rises to 5%, then flattens. The 5% threshold is 2.7x more conservative than statistical limit of 1.9%.

### 2.10. Recurrence analysis and Annotation

Heteroplasmic calls were grouped by position and substitution across the cohort. Positions called in one sample were classified as private. Each call was then cross-tabulated against homoplasmic variants (allele fraction ≥ 95%) in the cohort. Macrohaplogroup analysis was also done to avoid confusing a normal, ancient human variation (a lineage marker) with potential heteroplasmy. Calls at any of 165 lineage positions (32 from PhyloTree, 133 homoplasmic in ≥ 2 samples) were retained.

The variant calls related to Protein-coding regions were annotated with gene, codon, codon change, amino-acid change and consequence, using the vertebrate mitochondrial genetic code (translation table 2), as the amino acid translation codes in mitochondria slightly differ from genomic translation codes (Barrell et al., 1979). MT-ND6 was annotated on the light strand. For variant calls that span overlapping gene regions (MT-ATP8/MT-ATP6 and MT-ND4L/MT-ND4), the first gene in genomic order was reported.

**Table 2:** Mitochondrial coverage and callability across 47 samples. Quality-filtered depth uses base quality ≥ 20 and mapping quality ≥ 30.

| Metric | Mean | Median | Range |
| --- | --- | --- | --- |
| Depth, quality-filtered (×) | 433 | 226 | 9-3029 |
| Depth, MAPQ ≥ 30 (x) | 645 | 348 | 16-4513 |
| Breadth ≥ 50x (%) | 93.5 | 99.6 | 2.3-100.0 |
| Breadth ≥ 100x (%) | 85.6 | 97.5 | 0.0-100.0 |
| Breadth ≥ 500x (%) | 37.8 | 12.8 | 0.0-99.9 |
| Callable at 3% VAF (%) | 70.3 | 86.7 | 0.0-100.0 |
| Callable at 5% VAF (%) | 83.7 | 96.9 | 0.0-100.0 |
| Homoplasmic variants (n) | 29 | 31 | 0-52 |
| Heteroplasmic calls (n) | 3 | 2 | 0-10 |

Calls were also annotated against the gnomAD v3.1 mitochondrial reduced-annotations table, recording homoplasmic and heteroplasmic population frequencies, maximum observed heteroplasmy and gnomAD quality filters. Calls at positions flagged by gnomAD as artifact-prone or as NUMT-associated false-positive sites (common_low_heteroplasmy) were identified separately. ClinVar was not used as clinically relevant mitochondrial variants generally require high heteroplasmy to produce a phenotype.

### 2.11. Haplogroup assignment and lineage analysis

Macrohaplogroups were assigned with a marker-based classifier that runs offline, in place of HaploGrep3, which requires network access to PhyloTree. Eleven diagnostic positions were used (rCRS coordinates): five separating N from L3 and M (m.8701, 9540, 10398, 10873, 15301), four defining M (m.489, 10400, 14783, 15043) and two separating R from other N lineages (m.12705, 16223). The base at each position was read directly from the final chrM alignment (base quality ≥ 20, mapping quality ≥ 30). A marker was considered typed when at least 10 reads covered it and the major allele made up at least 80% of them; markers below this fraction were recorded as mixed. A sample was assigned only when at least three of the five N-block markers were typed and agreed; otherwise, it was reported as unassigned. Several markers per split were used, rather than one, so that assignment would tolerate the uneven coverage expected across a 9× to 3,029× depth range. Two or more mixed markers in a sample were taken to indicate possible contamination. Subclades were not assigned. To assess whether heteroplasmic calls reflected lineage variation, each call was cross-tabulated against the cohort’s homoplasmic variants. A call was classified as private if no sample carried that position homoplasmically. Calls at phylogenetically recurrent positions were flagged, not removed.

### 2.12. Data analysis and Visualisation

Figures were generated in Python 3.13.15 with matplotlib 3.10.0. All tabulation and summary statistics used the Python standard library; Spearman’s rank correlation (tie-corrected, with a normal-approximation two-sided p) and the hypergeometric tail probability were implemented directly rather than through an external statistics package. The workflow, configuration file, and container definition are available on GitHub at https://github.com/tomar-neeri/mtdna-heteroplasmy-host-reads and archived on Zenodo at https://doi.org/10.5281/zenodo.23034211. Variant calls and summary tables are provided as **Supplementary Data**. Software versions and analysis parameters are listed in **Supplementary Table S14.**

## 3. Results

### 3.1. Recovery of mitochondrial sequence from host reads

Across all 47 samples, the Mean depth was observed to be 433× (median 226×, range 9-3,029×) after quality filtering. Or 645× when counting all primary alignments at mapping quality ≥ 30. Breadth averaged 93.5% at ≥ 50× and 85.6% at ≥ 100×. The callable fraction was observed at 70.3% of positions cohort-wide at a 3% allele fraction (median 86.7%), rising to 83.7% at a 5% allele fraction. 20 samples reached ≥90% callability at 3%. Two samples (403, 407) fell below 50× mean depth and were excluded from heteroplasmy analysis, while retained for haplogroup typing. The results are summarised in **Table 2**

### 3.2. Removal of NUMT and Microbial reads contamination

Alignment against the human reference genome (GRCh38) located 115,302 of 3,908,080 recruited reads (2.95% pooled; per-sample mean 3.22%, range 1.53-6.35%) to nuclear sites, indicating NUMT contamination. chr1 was the predominant destination in 47 of 47 samples; reads also aligned to other chromosomes, but at very low levels **(Figure 1a-c).** The relocated fraction was independent of library size across a 211-fold range (ρ = −0.08, p = 0.58). Removing this 2.95% of reads removes 87.6% of heteroplasmic calls: without competitive realignment, the workflow returned 1,081 calls against 134 with it, and 79 calls above the 5% limit against 16 **(Supplementary Table S14).**

**Figure 1:**
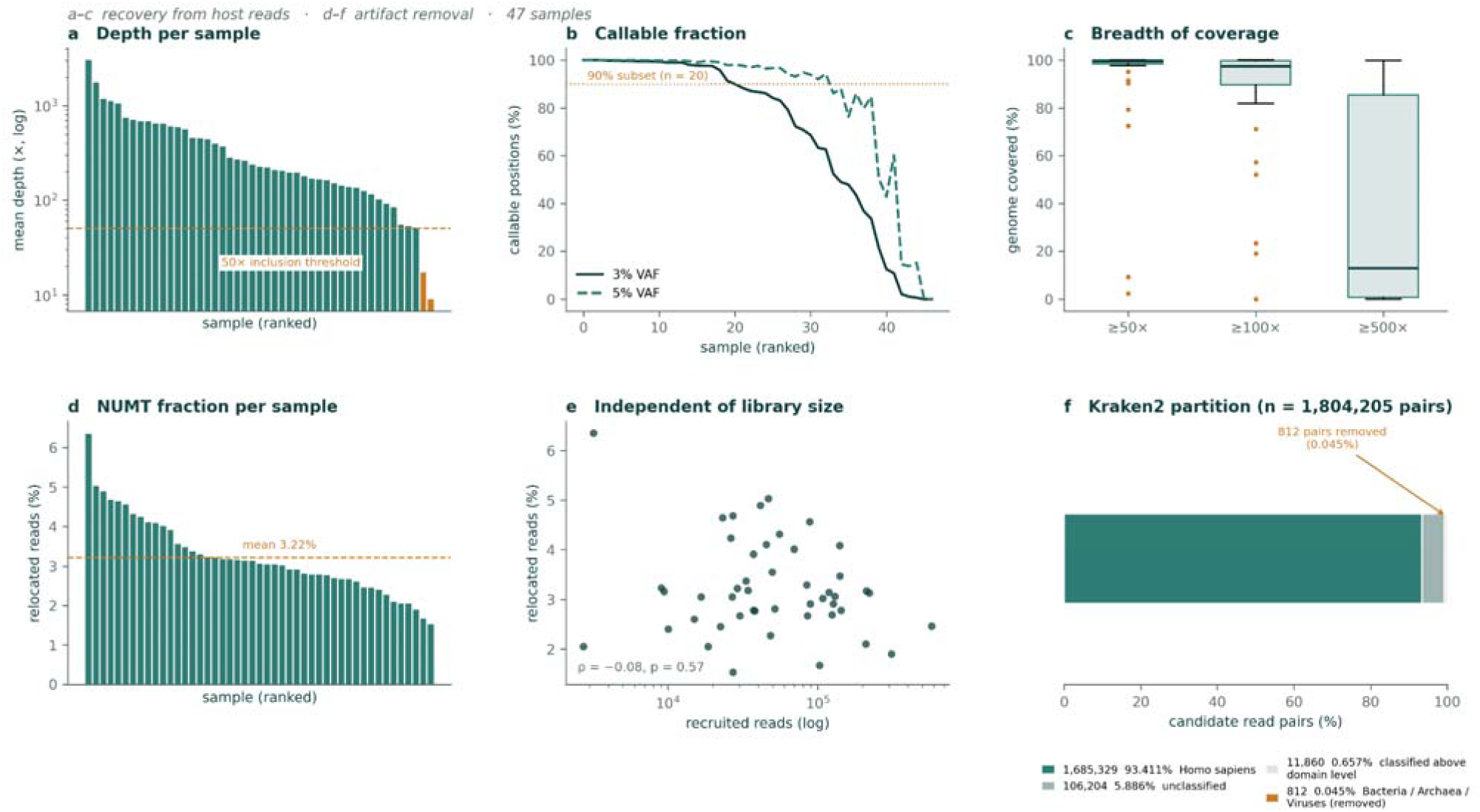
(a) Mean depth per sample, ranked; the two samples below the 50× threshold are shown in orange. (b) Callable fraction per sample at 3% and 5% VAF; the dotted line marks the 90% criterion, subset (n = 20). (c) Breadth of coverage at three depth thresholds. (d) Percentage of recruited reads relocating to nuclear loci on realignment against hg38, per sample; the dashed line is the sample group mean. (e) Relocated fraction of reads against number of recruited reads. (f) Taxonomic partition of the candidate read pairs entering Pass 2b; only reads assigned to Bacteria, Archaea or Viruses were removed.

Taxonomic classification using Kraken2 passed the 1,804,205 candidate read pairs entering Pass 2b as follows: 1,685,329 (93.4%) were classified as Homo sapiens, 106,204 (5.9%) were unclassified, 11,860 (0.7%) were classified only above domain level, and 812 (0.045%) fell beneath Bacteria, Archaea or Viruses and were removed. Bacterial assignments accounted for 811 of these; a single read pair was archaeal, and none was viral. No other eukaryotic taxon was detected. The maximum removal for any sample was 0.644%. **(Figure 1; d-f)** Because the Standard Kraken2 database contains the human reference genome, 93.4% of candidate reads were confidently classified rather than left unclassified; if the filter had retained only unclassified reads, it would have discarded all classified human reads. This means a loss of 1.69 million read pairs to remove 812. Microbial reads are therefore excluded at recruitment rather than at classification. A read reaches Pass 2b only if it first cross-mapped to rCRS. at mapping quality ≥ 30 as a proper pair. Classification then removed 0.045% of those that did, and Pass 2b using Kraken2 accordingly functions as a verifiable negative control rather than a necessary filter. The correct logic for removing reads mapping to non-human references is stored in the config.yaml file of the pipeline as kraken_exclude: “Bacteria, Archaea, Viruses”

### 3.3. Error model

Pooled fitting over 2,083,234 background observations gave α = 0.774, β = 3,902 and a mean error rate of 1.98 × 10⁻ (Q37.0), with overdispersion 2.56 × 10⁻ . All twelve substitution classes exceeded the 500-observation minimum and were fitted independently. Fitted rates spanned 12.8-fold, from T>A (7.02 × 10⁻ , Q31.5, 3.54× the global rate) to C>G (5.47 × 10⁻ , Q42.6, 0.28×). The distribution across classes is structured rather than random. Both directions of the A: T transversion were elevated (T>A Q31.5, A>T Q34.7) and both directions of the C:G transversion suppressed (G>C Q39.4, C>G Q42.6), a 6.2-fold difference between the two pair means. Transitions occupied a narrower range (Q36.1–38.2, 1.63-fold) close to the global rate. **(Figure 2)** and **Supplementary Table S5.**

**Figure 2:**
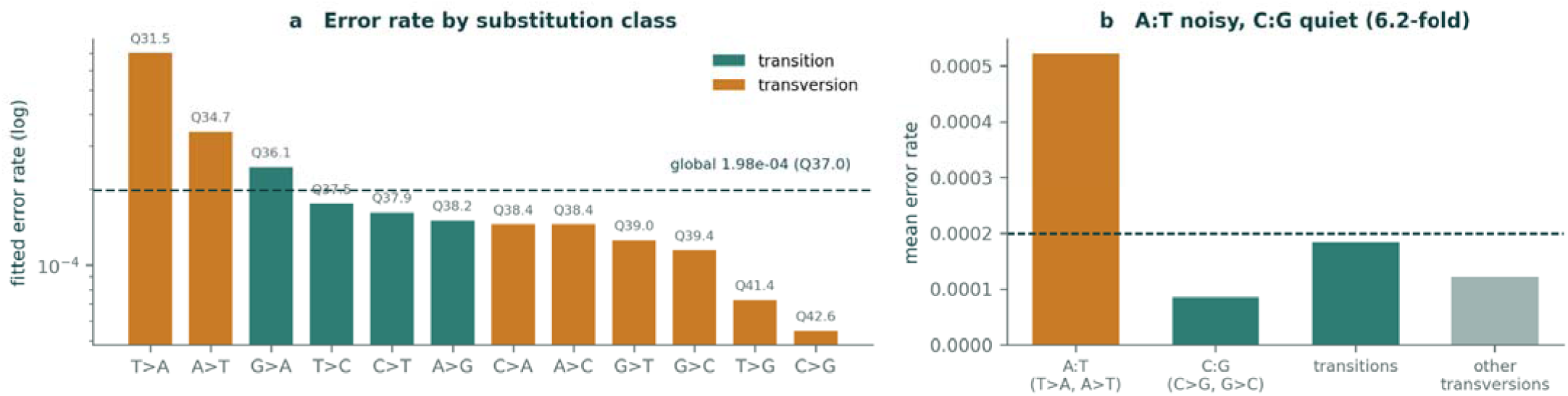
(a) Mean Error (log) rates by 12 substitution classes, with transition shown in green and transversions shown in orange. (b) Mean error rate across substitution classes, A:T showing 6.2x higher mean error rates.

### 3.5. Recurrence of Substitution and Haplogroup Composition

Thirteen of the 16 recurrent positions were transversions, ten of them T>A, and recurrence was concentrated at low allele fractions: no recurrent position had a median allele fraction above 4.07% **(Figure 3a, b)**. Two recurrent positions produced calls above the 5% threshold. m.16060G>A was called in 5 of 47 samples at 3.88–7.18%, and m.12310A>T in 5 samples at 1.18–5.26%. Neither is present in gnomAD v3.1, nor was it homoplasmic in any sample, and the carriers of each included samples from both major macrohaplogroups.

**Figure 3:**
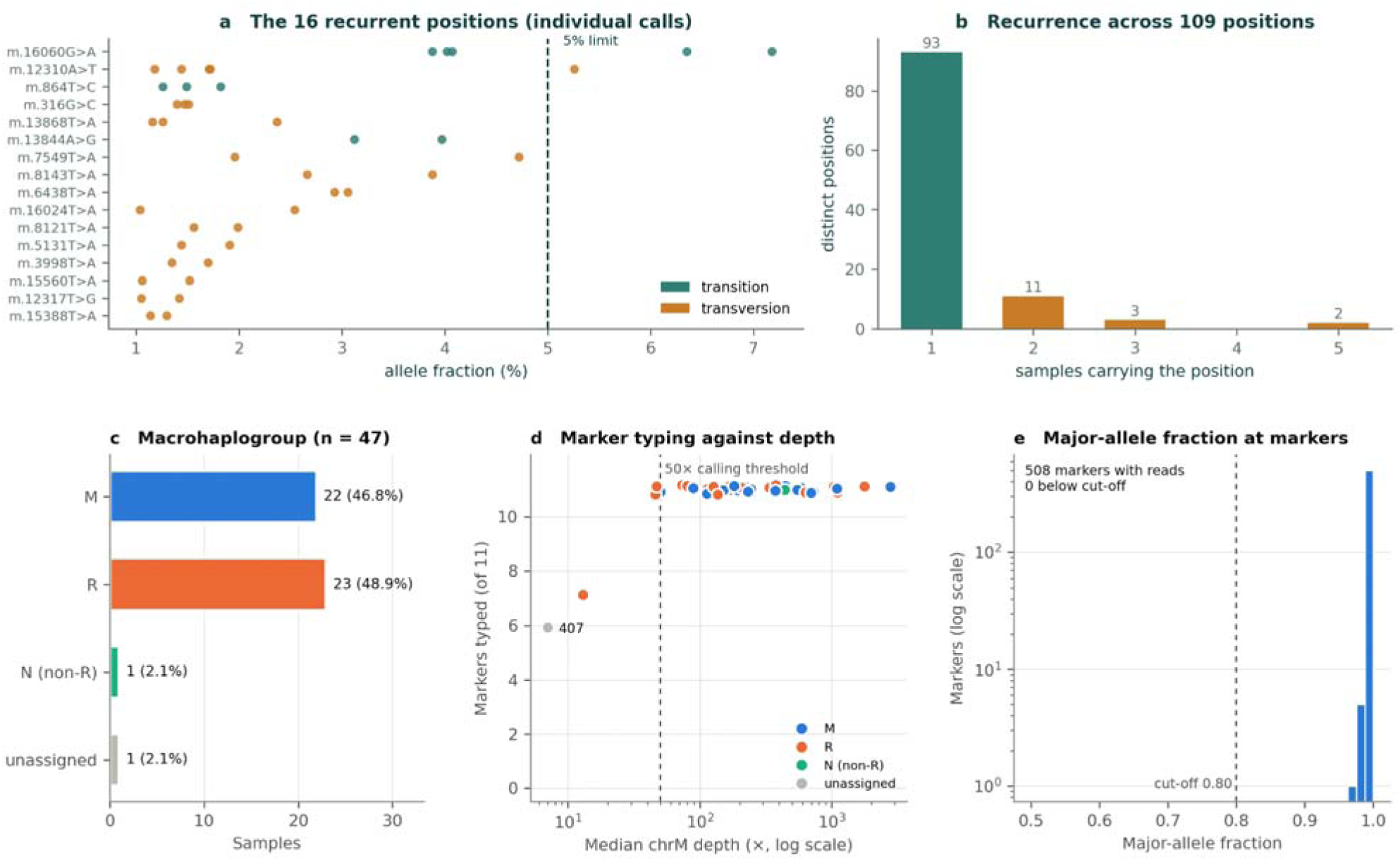
(a) Allele fraction of every call at the 16 positions called in more than one sample, coloured by substitution class; the dashed line marks the 5% threshold. (b) The 109 distinct positions by number of carrying samples; positions private to one sample are shown in blue, and recurrent positions in orange. (c) Macrohaplogroup assignment (n = 47). (d) Number of the 11 diagnostic markers typed (≥10 reads, major allele ≥80%) against median chrM depth; the dashed line marks the 50× calling threshold. All samples above it were typed at 11 of 11; the two samples below it were typed at 7 (403, assigned R) and 6 (407, unassigned). (e) Major-allele fraction at all 508 typed markers; none fell below the 0.80 cut-off that would indicate a mixture.

Macrohaplogroups were assigned to 46 of 47 samples: M 22 (46.8%), R 23 (48.9%) and N outside R 1 (2.1%) **(Figure 3c)**. All 11 diagnostic markers were typed in 45 samples. The two samples below the 50× calling threshold were typed at 7 (403) and 6 (407) markers, because typing requires only 10 reads per marker. Sample 403 produced no homoplasmic calls at its depth but showed reference bases at the typed markers and was assigned to R; sample 407 could not be assigned **(Figure 3d)**. All 508 typed markers had a major-allele fraction of at least 0.80 (lowest ≈0.97), so no sample showed the intermediate fractions expected of a mixture **(Figure 3e)**. The M/R balance is consistent with published Indian distributions.

### 3.6. Substitution Detection Limit

The substitution spectrum of the full call set generated only by the statistical model is not biologically plausible. Ti/Tv was 0.43 against 3.51 in gnomAD, with T>A alone accounting for 57 of 134 calls (42.5%) while C>T, the most common class in gnomAD, contributed one. Across 57 T>A calls, the maximum allele fraction was 4.72%, whereas transitions reached 90.81%. Real heteroplasmy occur clonally and spans the across the full range; an error process has a magnitude it cannot exceed. The 5% threshold was chosen based on the observed T>A maximum, so this probability quantifies the separation rather than being tested on independent data. Raising the threshold removed the error component. T>A calls fell from 57 at 1% to 12 at 2%, 5 at 3% and none at or above 5%, while Ti/Tv rose from 0.43 to 2.20 and then plateaued (2.00 at 7%, 2.33 at 10%). No further improvement was obtained above 5%. Ninety-one of 134 calls (68%) lay in the 1-2% band, where Ti/Tv was 0.43. (**Figure 4; a-c)** and **Supplementary Table S6**

**Figure 4:**
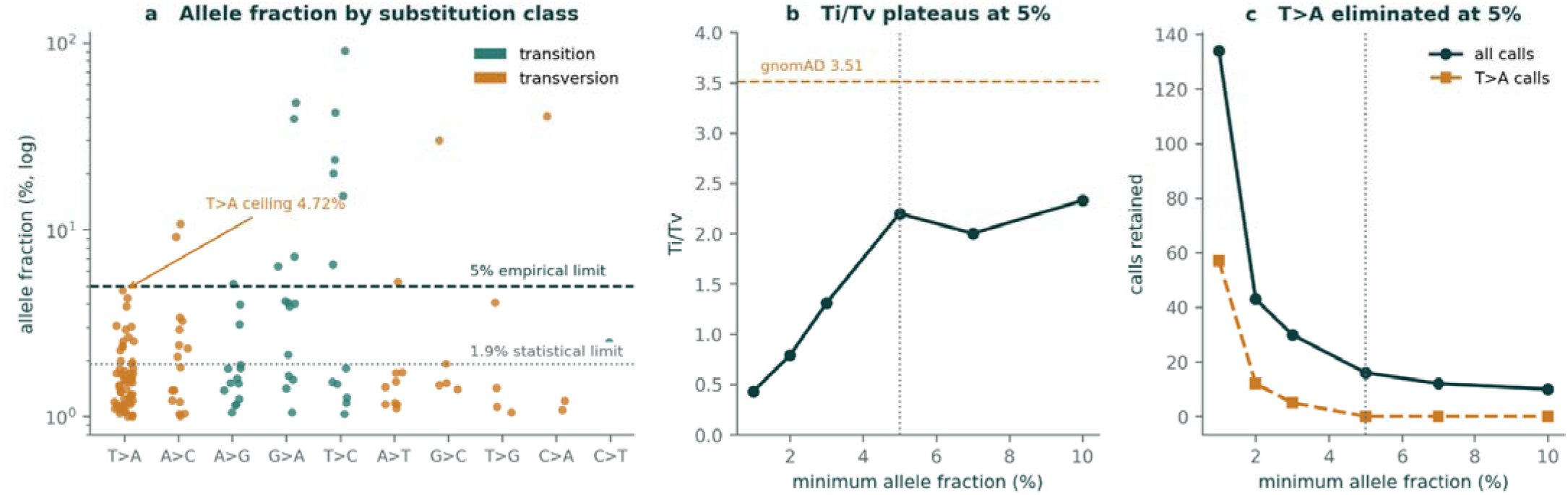
(a) Allele fraction % (log) by substitution class (Transversion/Transition) crossing the statistical (1.9%) and empirical (5%) thresholds. (b) Ti/Tv ratio at different allele fraction % thresholds; the Ti/Tv ratio flattened at 5% allele fraction (c). T>A calls at different allele fraction % thresholds. T>A calls were reduced to zero at an allele fraction % threshold of 5%, depicted by a vertical dotted line.

### 3.7. Final Heteroplasmy Calls

Our analysis detected 134 heteroplasmic calls across 40 of 47 samples (mean 2.85 per sample, maximum 10), of which 16 clear the 5% threshold and 10 clear of gnomAD’s 10% threshold. Calls were predominantly in protein-coding sequences (102 calls, 35 samples, median 1.54%) and the control region (23 calls, 19 samples, median 3.38%), with 8 in rRNA genes and 1 in tRNA genes. Of coding calls, 62 were missense, 16 synonymous and 9 nonsense.

Against gnomAD, 79 calls were novel, 24 rare, 18 previously observed at heteroplasmy and 10 known polymorphisms. Three calls, all m.316G>C, in three samples at 1.40-1.51%, fell at a position gnomAD flags as artifact-prone; m.316 lies adjacent to the masked poly-C tract at m.302– 315, and all three fall below the 5% limit.

Three calls (2.2%) fell at flagged lineage positions: m.16093T>C (90.8%), m.204T>C (20.1%) and m.16183A>C (10.8%), all in the control region and all above the 5% limit. Cross-tabulation against the cohort homoplasmic landscape classified 130 of 134 calls as private, with two shared across macrohaplogroups (m.16093T>C, m.204T>C), one lineage-associated within the caller’s own macrohaplogroup (m.8084A>C, 1.22%), and one shared with a different macrohaplogroup only (m.15530T>A, 1.13%). **(Figure 5; a-d)** and **Supplementary Table S2**

**Figure 5:**
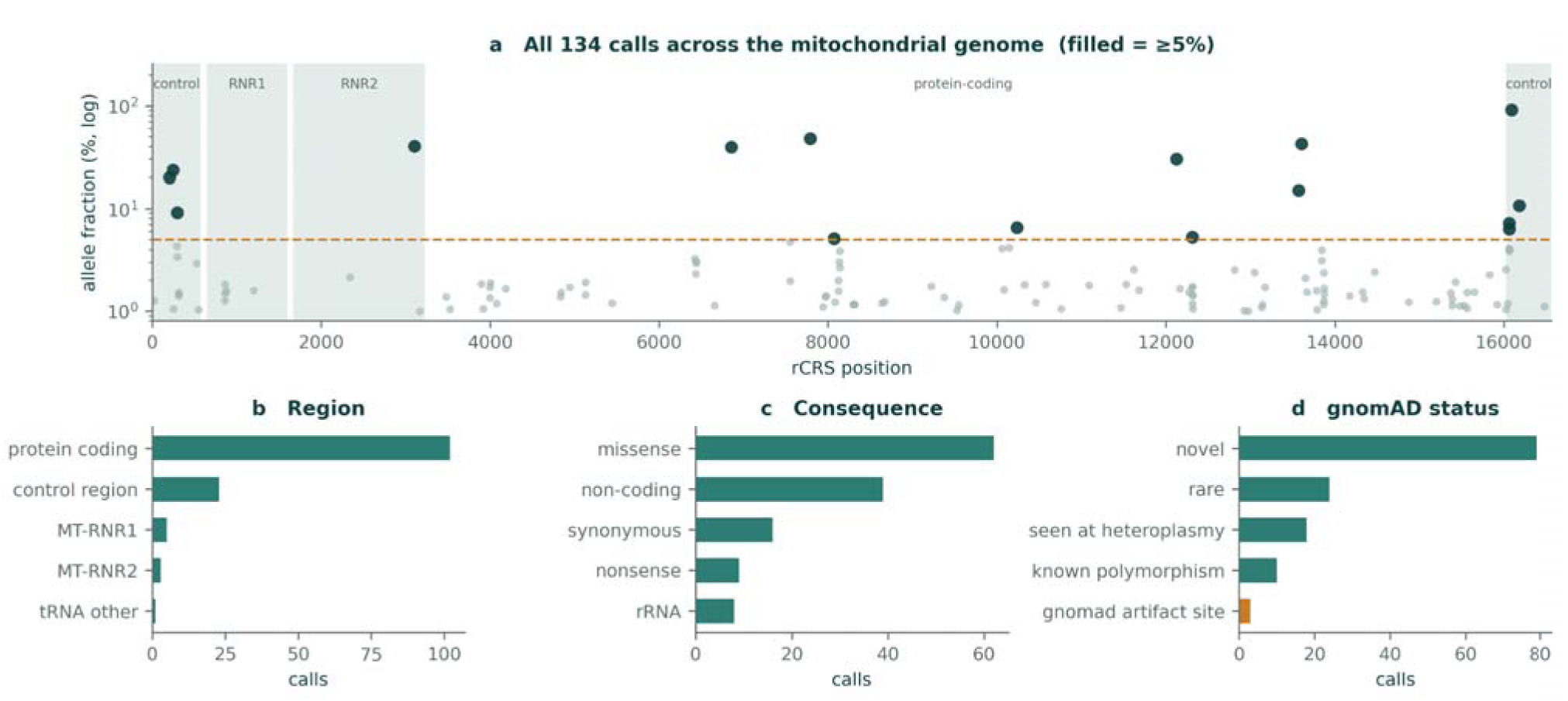
(a) Summary of 134 heteroplasmy calls mappled across the length of mitochondrial reference sequence, with the region-specific annotation, dotted line depicting the 5% threshold. (b) genomic regions mapped against the number of calls. (c) Mutation consequences mapped against the number of calls. (d) gnomAD annotations of calls mapped against the numbers.

## Discussion

Through this study, we sought to establish the utility of otherwise discarded human host reads from shotgun metagenomic data for mitochondrial heteroplasmy profiling. This approach is conceptually supported by the work of (Griffin et al., 2014) who demonstrated that off-target reads from whole-exome sequencing could be used to accurately detect pathogenic point mutations in mitochondrial DNA. However, our approach differs from previous studies in that we investigate whether human reads routinely discarded during shotgun metagenomic analysis can be repurposed for quantitative mitochondrial heteroplasmy detection. Our study not only focuses on the feasibility of detection but also defines the analytical limits within which such detection remains reliable.

Previous studies have used mitochondrial reads recovered from metagenomic datasets for applications such as haplogroup assignment (Ma et al., 2014) sample re-identification, and contamination assessment (Tomofuji et al., 2023). Human mitochondrial haplogroups have also been recovered from urban sewage metagenomes (Pipek et al., 2019), and from shotgun-sequenced palaeofaeces (Wibowo et al., 2021) However, to our knowledge, none has explored the feasibility of detecting mitochondrial heteroplasmy in human URT shotgun metagenomes and evaluated its associated error structure. Thus, our study extends the utility of host-derived reads in shotgun metagenomics from population and sample-level characterisation toward quantitative mitochondrial variant profiling, having also defined the analytical constraints of this approach.

We have shown that all the samples have shown complete mitochondrial genome coverage at mean 433x depth. This study has established that recovery of complete mitochondrial genomes is not a constraint from URT shotgun metagenomics data. The constraints come from other sources, the primary constraints were the cross mapping of NUMTs with the mitochondrial DNA and sequencing errors that are selective to certain substitution types and genomic locations at the mitochondrial genome. The pipeline addresses these constraints and enables calling of mitochondrial heteroplasmy above 5% VAF.

Host reads that metagenomic workflows routinely discarded could therefore add a host-genetic layer to microbiome studies without additional sequencing. This may be particularly relevant to tumour microbiome studies, where host mitochondrial variation is a potential confounder of, or modifier of, associations between microbial communities and disease. Heteroplasmy measured in this way could allow such studies to account for host mitochondrial state when interpreting microbial associations. The present work defines the analytical limits within which such studies could use host reads.

A major result of our work is the definition of a reliable detection limit for the proposed methodology; we have observed that statistical models alone cannot establish a reliable detection threshold for heteroplasmy under given data conditions. The variant call sub-set only from the statistical model had a Ti/Tv of 0.43 against 3.51 in gnomAD. T>A accounted for 42.5% of calls and C>T for one. These preliminary results are incompatible with established mitochondrial mutational biology. Duplex sequencing across tissues shows somatic mtDNA mutations to be strongly transition-biased, with few transversions (Sanchez-Contreras et al., 2023) whereas this was not the case with the initial variant call set. The allele-fraction ceiling is the stronger evidence. T>A never exceeded 4.72% in 57 calls, whereas transitions reached 90.81%. Raising the threshold removed T>A entirely at 5%, and Ti/Tv rose to 2.20 and then plateaued. This specific failure of the error model is largely due to a structural reason; the background is defined as positions below 0.5% VAF. An error process operating above 0.5% level at scattered private positions therefore passes the significance test. Our threshold of 5% VAF converges with reported threshold ranges but from a different route; the original gnomAD paper reported that after finding many false positive calls below 10% VAF, it excluded samples with mtDNA copy number under 50 and reported only variants at or above 10% (Laricchia et al., 2022) It is important to note that the gnomAD thresholds are cohort-specific and were tested on a larger cohort of 56,434 individuals than in our study (47), and therefore are not directly comparable. The original paper also states that further releases of gnomAD could reduce the threshold to 5% VAF. The thresholds are not directly transferable, however. gnomAD’s cutoffs were set for blood-derived whole genomes to control NUMT false positives that scale with mitochondrial DNA copy number, which is not measured here. Our threshold was derived from metagenomic URT swab data to control a substitution-specific error that persisted after NUMT removal. The agreement is therefore between independent lines of evidence reaching a similar allele fraction.

The 5% threshold alone does not certify every call. Four positions warrant particular caution. m.16060G>A and m.12310A>T are recurrent sites. m.316 is flagged artifact-prone by gnomAD and lies next to the m.302-315 poly-C tract, and m.16183 lies immediately next to the m.16184-16193 tract. gnomAD’s own coverage profile shows depth dips at positions 303-315.

We have characterised and tried to rule out the T>A artifact from the variant calls but the reason behind its higher presence could not be ascertained with the available data. With T>A is not having a sequence context (1.2–2.2-fold variation across trinucleotides), not homopolymers (39% of calls flanked by T against 43% of background), and not position (46 distinct sites spanning m.22– 16,024). The error model adds one constraint: both directions of the A:T transversion are elevated (Q31.5 and Q34.7), while both directions of the C:G transversion are suppressed (Q42.6 and Q39.4), as shown in **Supplementary Figure S1**.

The macrohaplogroup distribution among the 46 (n=46) assigned samples (M 22, 47.8%; R 23, 50.0%; N outside R 1, 2.2%) is consistent with South Asian populations, although M falls at the lower end of reported values. Earlier studies found that macrohaplogroup M was present in about 60% of Indian individuals (Metspalu et al., 2004). and it averaged 70% across 26 tribal groups (Chandrasekar et al., 2009) Within N, haplogroup R and its sub-haplogroups, including U, predominate in India. An urban, largely non-tribal cohort would be expected to sit nearer to these numbers. The reference rCRS genome against which such data are interpreted are largely dependent on European lineages. gnomAD v3.1 is 58% European and about 5% Asian by ancestry (3% South Asian), and the rCRS itself belongs to the European haplogroup H (Laricchia et al., 2022) Due to this, two consequences could follow. First, population heteroplasmy baselines are drawn largely from European haplogroups. Second, M-lineage samples differ from the reference at more homoplasmic positions than R-lineage samples, which could affect alignment and calling.

Three failure modes were observed while analysing each, which produce well-formed output but a zero-exit status. The most significant one occurs during host depletion; the Standard Kraken2 database contains the human genome, so genuine mitochondrial reads are confidently classified as Homo sapiens. Retaining only unclassified output, the conventional approach to microbial depletion, and the default behaviour of “unclassified-out” discards 92% of the target mitochondrial data. Depletion by taxonomic assignment and depletion by classification status are not interchangeable operations when the database contains the host. The other two were an unbounded beta-binomial concentration, which drives α + β toward 10¹ until the survival function returns exactly zero and the significance filter silently disables itself, and R1/R2 desynchronisation from a coordinate-sorted BAM, after which bwa mem emits no alignments and exits cleanly. We have elaborated on these failure modes in the pipeline’s README.

### Limitations

At this stage, external validation of the pipeline using open-source data is not possible, as all public databases, such as the NCBI Sequence Read Archive (SRA), perform host removal at submission, which limits the applicability of this pipeline to public data. Since the samples from the control arm of a retrospective parent study were used, and there were no replicate samples. Therefore, the replicate comparisons could not be performed. A replicate comparison would be performed in future studies, as it is the most direct test of whether a sub-5% call is reproducible. Certain Confounders should also be considered while interpreting the results; heteroplasmy calls varied widely and correlated with depth. Although heteroplasmic variants arise largely somatically and rise sharply after 70 years of age (Gupta et al., 2023) But in this group, the median age was 28 years (IQR 25-40, range 22-54); therefore, no age-based analysis was done.

We have limited the assignments to macrohaplogroups while accounting for low-sequencing-depth samples; however, a deeper phylogenetic analysis should be explored in further studies that generate richer data. The cohort represents a single tissue, population and sequencing pipeline, so the thresholds reported here may need re-optimisation in other settings.

## Conclusion

Host reads from shotgun metagenomes support complete mitochondrial recovery and haplogroup typing in nearly every sample, and heteroplasmy detection in most of the samples. The dominant artifact is of nuclear origin, not microbial. What remains after competitive realignment is a T>A-dominated error component that background-fitted models cannot capture due to its higher frequency. Heteroplasmy calls from these data are reliable at approximately 5% allele fraction and above. The findings demonstrate the potential utility of metagenomic host reads alongside whole-genome resources.

## Data availability statement

The workflow, configuration file, and container definition are available on GitHub (https://github.com/tomar-neeri/mtdna-heteroplasmy-host-reads) and archived on Zenodo (https://doi.org/10.5281/zenodo.23034211). Heteroplasmic variant calls and summary statistics are provided as **Supplementary Data**. Host-depleted metagenomic reads are available under BioProject: PRJNA1478410

## Author declaration

The authors assure that the research has complied with all ethical guidelines and has received approval from the Institutional Ethics Committee for Research on Human Subjects (IEC) of CSIR-NEERI, Nagpur-20, India **(Reference No: Eth.Com./001/IEC/02/2023)**. Necessary consent from patients/participants has been obtained, and relevant institutional documentation has been archived. This manuscript is approved by the institutional Knowledge Resource Center (KRC) of CSIR-NEERI, bearing KRC No. CSIR-NEERI/KRC/2026/OCT/EEPM/1

## Confidentiality declaration

Sample IDs are masked IDs and cannot be traced to participant details. The precise ages of the participants are masked, and non-overlapping age ranges are used.

## Author contribution statement

SST and KK have contributed equally to the conceptualization, experimentation, and data analysis of this study.

## Data Availability

The workflow, configuration file, and container definition are available on GitHub (https://github.com/tomar-neeri/mtdna-heteroplasmy-host-reads) and archived on Zenodo (https://doi.org/10.5281/zenodo.23034211). Heteroplasmic variant calls and summary statistics are provided as Supplementary Data. Host-depleted metagenomic reads are available under BioProject: PRJNA1478410

https://doi.org/10.5281/zenodo.23034211

## Acknowledgement

The authors are thankful to CSIR-NEERI for providing funds under project OLP-57 (March 2023 -April 2024) for conducting this study.

---

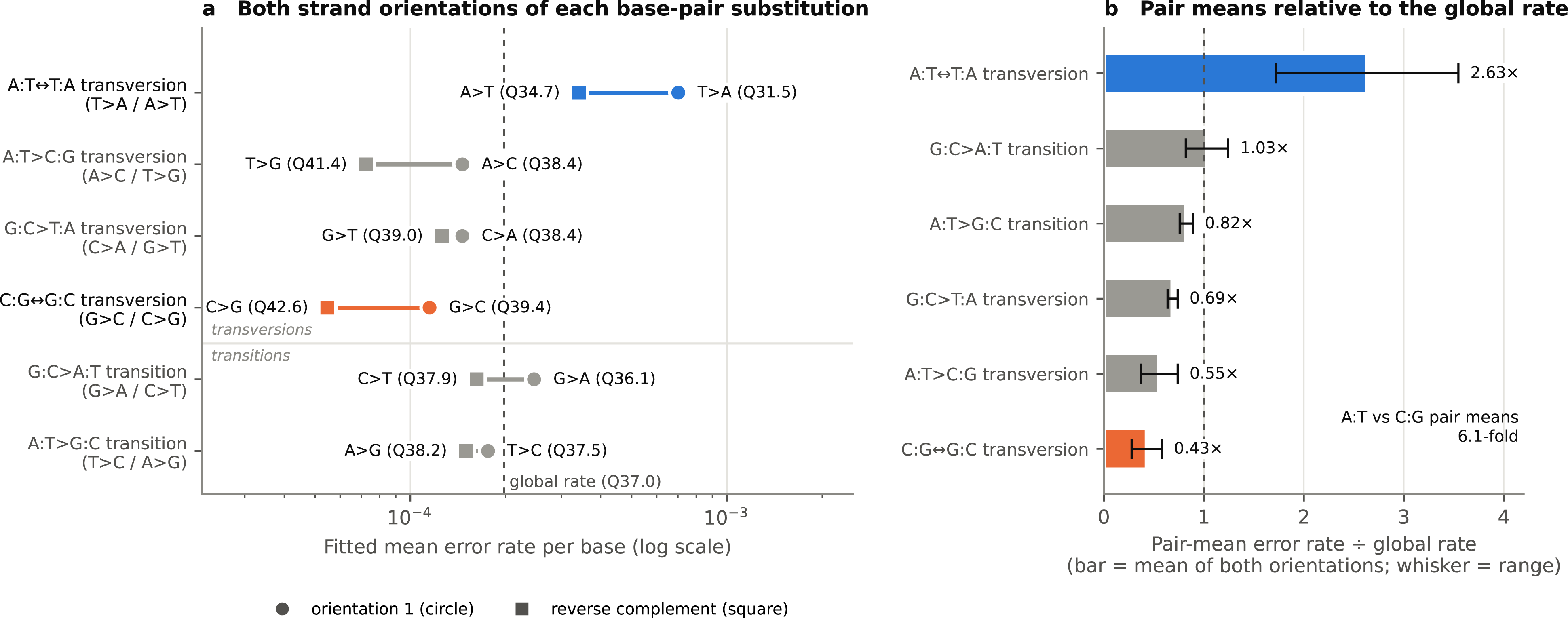

