## Supplementary Table for "Repurposing Shotgun Metagenomic Human Host Reads for Mitochondrial Heteroplasmy Detection"

&

^2^ Academy of Scientific and Innovative Research (AcSIR), Ghaziabad, UP, India.

*Corresponding author: Correspondence to Krishna Khairnar

**Table S1.** Per-sample mitochondrial coverage, callability and variant counts

**Table S2.** NUMT relocation on competitive realignment to GRCh38

**Table S3.** Kraken2 taxonomic partition of candidate read pairs (total)

**Table S4.** Per-sample Kraken2 classification and removal

**Table S5.** Pooled substitution-specific beta-binomial error model

**Table S6.** Transition/transversion ratio and T>A calls across allele-fraction thresholds

**Table S7.** All heteroplasmic calls with functional, population and lineage annotation (n = 134)

**Table S8.** Positions called heteroplasmic in more than one sample (n = 16)

**Table S9.** Heteroplasmic calls at lineage positions or shared with homoplasmic carriers in the samples

**Table S10.** Distribution of heteroplasmic calls by genomic region

**Table S11.** Macrohaplogroup assignment

**Table S12.** Allele calls at the eleven diagnostic macrohaplogroup markers

**Table S13.** Software versions and analysis parameters

1. **Sample identifiers, the three-digit study codes, could not be mapped directly to the participant details.**
2. **VAF (variant allele fraction)**
3. **All coordinates refer to the rCRS (NC_012920.1)**

### **Supplementary Table S1. Per-sample mitochondrial coverage, callability and variant counts**

| **Sample** | **Mean depth (QF)** | **Median depth (QF)** | **Mean depth (MAPQ≥30)** | **Breadth ≥50× (%)** | **Breadth ≥100× (%)** | **Breadth ≥500× (%)** | **Uncovered bases** | **Callable 3% VAF (%)** | **Callable 5% VAF (%)** | **Callable ≥90% at 3%** | **Homoplasmic (n)** | **Heteroplasmic candidates (n)** | **Heteroplasmic calls (n)** | **Heteroplasmy analysis** |
| --- | --- | --- | --- | --- | --- | --- | --- | --- | --- | --- | --- | --- | --- | --- |
| 387 | 141 | 125 | 209 | 98.4 | 89.9 | 2.5 | 0 | 43.5 | 86.1 | No | 25 | 2 | 1 | Yes |
| 388 | 236 | 226 | 372 | 99.7 | 97.4 | 19.0 | 0 | 87.1 | 96.9 | No | 33 | 3 | 2 | Yes |
| 389 | 710 | 656 | 1021 | 100.0 | 100.0 | 91.2 | 0 | 99.7 | 100.0 | Yes | 38 | 14 | 4 | Yes |
| 390 | 261 | 246 | 407 | 99.6 | 98.8 | 24.3 | 0 | 91.3 | 97.9 | Yes | 34 | 11 | 7 | Yes |
| 391 | 179 | 171 | 264 | 99.5 | 94.8 | 3.2 | 0 | 72.3 | 93.1 | No | 23 | 1 | 1 | Yes |
| 392 | 226 | 214 | 345 | 99.5 | 97.5 | 12.8 | 0 | 84.2 | 96.7 | No | 29 | 6 | 4 | Yes |
| 393 | 1055 | 1036 | 1615 | 100.0 | 100.0 | 97.4 | 0 | 99.8 | 100.0 | Yes | 30 | 13 | 2 | Yes |
| 394 | 644 | 646 | 1036 | 99.9 | 99.7 | 91.4 | 0 | 99.0 | 99.7 | Yes | 40 | 6 | 3 | Yes |
| 395 | 561 | 511 | 844 | 100.0 | 99.8 | 84.2 | 0 | 99.0 | 99.7 | Yes | 30 | 12 | 4 | Yes |
| 396 | 151 | 151 | 225 | 98.5 | 94.0 | 0.0 | 0 | 63.2 | 92.0 | No | 9 | 2 | 2 | Yes |
| 397 | 165 | 162 | 251 | 99.5 | 96.1 | 0.9 | 0 | 70.8 | 94.9 | No | 33 | 3 | 3 | Yes |
| 398 | 221 | 211 | 348 | 99.3 | 97.2 | 12.8 | 0 | 86.2 | 96.5 | No | 39 | 7 | 6 | Yes |
| 399 | 744 | 728 | 1112 | 100.0 | 99.9 | 93.6 | 0 | 99.5 | 99.9 | Yes | 30 | 12 | 4 | Yes |
| 400 | 135 | 132 | 204 | 98.5 | 91.6 | 0.6 | 0 | 48.9 | 88.0 | No | 30 | 2 | 2 | Yes |
| 401 | 115 | 115 | 175 | 97.8 | 88.6 | 0.0 | 0 | 33.6 | 84.7 | No | 34 | 0 | 0 | Yes |
| 402 | 1114 | 1104 | 1700 | 100.0 | 100.0 | 98.2 | 0 | 99.8 | 100.0 | Yes | 38 | 14 | 2 | Yes |
| 403 | 17 | 13 | 22 | 9.3 | 0.0 | 0.0 | 0 | 0.0 | 0.0 | No | 0 | 0 | 0 | No (<50×) |
| 404 | 84 | 73 | 119 | 91.6 | 52.0 | 0.1 | 0 | 12.4 | 42.8 | No | 27 | 0 | 0 | Yes |
| 405 | 650 | 582 | 972 | 100.0 | 99.9 | 86.8 | 0 | 99.4 | 99.9 | Yes | 25 | 10 | 2 | Yes |
| 406 | 195 | 169 | 290 | 99.0 | 95.7 | 9.9 | 0 | 68.7 | 93.9 | No | 35 | 1 | 0 | Yes |
| 407 | 9 | 7 | 16 | 2.3 | 0.0 | 0.0 | 0 | 0.0 | 0.0 | No | 0 | 0 | 0 | No (<50×) |
| 408 | 394 | 337 | 651 | 99.7 | 99.2 | 61.7 | 0 | 95.8 | 99.1 | Yes | 38 | 3 | 1 | Yes |
| 409 | 102 | 80 | 147 | 90.3 | 57.2 | 1.6 | 0 | 21.3 | 50.4 | No | 21 | 4 | 2 | Yes |
| 410 | 595 | 588 | 912 | 99.9 | 99.7 | 87.8 | 0 | 99.0 | 99.8 | Yes | 35 | 13 | 6 | Yes |
| 411 | 168 | 150 | 239 | 99.1 | 94.7 | 5.0 | 0 | 62.7 | 94.3 | No | 30 | 6 | 5 | Yes |
| 412 | 125 | 113 | 176 | 98.6 | 87.0 | 0.4 | 0 | 36.8 | 79.8 | No | 29 | 3 | 2 | Yes |
| 413 | 204 | 194 | 293 | 99.6 | 97.3 | 4.8 | 0 | 83.0 | 96.7 | No | 36 | 4 | 3 | Yes |
| 414 | 1179 | 1091 | 1681 | 100.0 | 100.0 | 97.4 | 0 | 100.0 | 100.0 | Yes | 37 | 7 | 1 | Yes |
| 415 | 454 | 444 | 658 | 99.9 | 99.1 | 72.2 | 0 | 97.8 | 99.1 | Yes | 38 | 23 | 10 | Yes |
| 416 | 369 | 376 | 537 | 99.9 | 99.5 | 56.6 | 0 | 97.6 | 99.4 | Yes | 34 | 10 | 2 | Yes |
| 417 | 453 | 441 | 698 | 99.9 | 99.6 | 75.5 | 0 | 98.0 | 99.5 | Yes | 34 | 14 | 8 | Yes |
| 418 | 1763 | 1770 | 2498 | 100.0 | 100.0 | 99.3 | 0 | 100.0 | 100.0 | Yes | 31 | 11 | 2 | Yes |
| 419 | 55 | 50 | 80 | 79.4 | 23.4 | 0.0 | 0 | 2.2 | 14.4 | No | 11 | 1 | 1 | Yes |
| 420 | 162 | 127 | 223 | 98.0 | 81.9 | 6.1 | 0 | 47.9 | 76.3 | No | 31 | 12 | 8 | Yes |
| 421 | 280 | 244 | 405 | 99.7 | 98.5 | 23.0 | 0 | 90.0 | 98.0 | No | 32 | 8 | 5 | Yes |
| 422 | 138 | 136 | 201 | 97.7 | 89.4 | 0.0 | 0 | 52.3 | 86.1 | No | 29 | 0 | 0 | Yes |
| 423 | 208 | 207 | 299 | 99.8 | 97.9 | 3.4 | 0 | 86.7 | 97.6 | No | 27 | 7 | 5 | Yes |
| 424 | 437 | 373 | 663 | 99.8 | 99.5 | 58.9 | 0 | 97.6 | 99.5 | Yes | 37 | 7 | 2 | Yes |
| 425 | 3029 | 2774 | 4513 | 100.0 | 100.0 | 99.9 | 0 | 100.0 | 100.0 | Yes | 32 | 13 | 2 | Yes |
| 426 | 91 | 89 | 132 | 95.2 | 71.2 | 0.0 | 0 | 10.7 | 60.1 | No | 26 | 1 | 1 | Yes |
| 427 | 51 | 46 | 75 | 72.4 | 23.3 | 0.0 | 0 | 1.1 | 13.7 | No | 19 | 1 | 1 | Yes |
| 428 | 606 | 550 | 840 | 100.0 | 100.0 | 76.9 | 0 | 99.5 | 100.0 | Yes | 52 | 19 | 5 | Yes |
| 429 | 53 | 47 | 73 | 72.6 | 18.9 | 0.0 | 0 | 0.6 | 15.1 | No | 16 | 0 | 0 | Yes |
| 431 | 682 | 633 | 1065 | 100.0 | 99.8 | 91.7 | 0 | 99.3 | 99.8 | Yes | 34 | 6 | 1 | Yes |
| 432 | 268 | 231 | 389 | 99.7 | 98.3 | 23.3 | 0 | 88.4 | 97.7 | No | 32 | 10 | 5 | Yes |
| 433 | 195 | 181 | 300 | 99.2 | 95.9 | 8.4 | 0 | 79.2 | 94.1 | No | 33 | 5 | 3 | Yes |
| 434 | 688 | 698 | 999 | 100.0 | 100.0 | 92.6 | 0 | 99.7 | 100.0 | Yes | 29 | 13 | 4 | Yes |

QF (quality-filtered depth; base quality ≥20, mapping quality ≥30) was used for calling. Mean depth (MAPQ≥30) counts all primary alignments at mapping quality ≥30.

Callable fraction at VAF f is the proportion of the 16,569 positions with depth ≥4/f.

Heteroplasmic candidates are positions with depth ≥50, ≥4 alternate reads and VAF between 1% and 95%, before the significance (including panel-of-normals) strand-bias filters

Heteroplasmic calls are those passing all filters. Samples 403 and 407 fell below 50× mean depth and were excluded from heteroplasmy analysis but retained for haplogroup typing.

Twenty samples reached ≥90% callability at 3% VAF.

### **Supplementary Table S2. NUMT relocation on alignment to GRCh38**

| **Sample** | **Recruited reads (Pass 1)** | **Retained on chrM (Pass 2)** | **Relocated to nuclear loci** | **Relocated (%)** | **Principal nuclear destination** |
| --- | --- | --- | --- | --- | --- |
| 387 | 26,944 | 26,121 | 823 | 3.05 | chr1 |
| 388 | 49,777 | 48,010 | 1,767 | 3.55 | chr1 |
| 389 | 130,628 | 126,631 | 3,997 | 3.06 | chr1 |
| 390 | 55,706 | 53,305 | 2,401 | 4.31 | chr1 |
| 391 | 34,338 | 33,247 | 1,091 | 3.18 | chr1 |
| 392 | 45,486 | 43,622 | 1,864 | 4.10 | chr1 |
| 393 | 211,660 | 204,947 | 6,713 | 3.17 | chr1 |
| 394 | 140,666 | 134,922 | 5,744 | 4.08 | chr1 |
| 395 | 108,466 | 105,187 | 3,279 | 3.02 | chr1 |
| 396 | 29,128 | 28,191 | 937 | 3.22 | chr1 |
| 397 | 33,315 | 32,191 | 1,124 | 3.37 | chr1 |
| 398 | 46,916 | 44,556 | 2,360 | 5.03 | chr1 |
| 399 | 143,486 | 139,495 | 3,991 | 2.78 | chr1 |
| 400 | 27,226 | 25,952 | 1,274 | 4.68 | chr1 |
| 401 | 23,250 | 22,172 | 1,078 | 4.64 | chr1 |
| 402 | 221,519 | 214,587 | 6,932 | 3.13 | chr1 |
| 403 | 2,738 | 2,682 | 56 | 2.05 | chr1 |
| 404 | 15,050 | 14,659 | 391 | 2.60 | chr1 |
| 405 | 124,954 | 121,592 | 3,362 | 2.69 | chr1 |
| 406 | 37,310 | 35,851 | 1,459 | 3.91 | chr1 |
| 407 | 3,198 | 2,995 | 203 | 6.35 | chr1 |
| 408 | 88,667 | 84,622 | 4,045 | 4.56 | chr1 |
| 409 | 18,610 | 18,229 | 381 | 2.05 | chr1 |
| 410 | 119,248 | 115,503 | 3,745 | 3.14 | chr1 |
| 411 | 30,304 | 29,494 | 810 | 2.67 | chr1 |
| 412 | 22,486 | 21,936 | 550 | 2.45 | chr1 |
| 413 | 37,378 | 36,338 | 1,040 | 2.78 | chr1 |
| 414 | 210,078 | 205,663 | 4,415 | 2.10 | chr1 |
| 415 | 84,842 | 82,053 | 2,789 | 3.29 | chr1 |
| 416 | 69,814 | 67,014 | 2,800 | 4.01 | chr1 |
| 417 | 89,480 | 86,877 | 2,603 | 2.91 | chr1 |
| 418 | 311,088 | 305,167 | 5,921 | 1.90 | chr1 |
| 419 | 10,096 | 9,854 | 242 | 2.40 | chr1 |
| 420 | 27,254 | 26,838 | 416 | 1.53 | chr1 |
| 421 | 51,786 | 50,329 | 1,457 | 2.81 | chr1 |
| 422 | 26,386 | 25,270 | 1,116 | 4.23 | chr1 |
| 423 | 38,132 | 37,074 | 1,058 | 2.77 | chr1 |
| 424 | 85,762 | 83,473 | 2,289 | 2.67 | chr1 |
| 425 | 577,931 | 563,710 | 14,221 | 2.46 | chr1 |
| 426 | 16,714 | 16,205 | 509 | 3.05 | chr1 |
| 427 | 9,452 | 9,153 | 299 | 3.16 | chr1 |
| 428 | 103,216 | 101,497 | 1,719 | 1.67 | chr1 |
| 429 | 9,064 | 8,771 | 293 | 3.23 | chr1 |
| 431 | 141,075 | 136,180 | 4,895 | 3.47 | chr1 |
| 432 | 48,432 | 47,332 | 1,100 | 2.27 | chr1 |
| 433 | 41,508 | 39,478 | 2,030 | 4.89 | chr1 |
| 434 | 127,516 | 123,803 | 3,713 | 2.91 | chr1 |
| **Total** | **3,908,080** | **3,792,778** | **115,302** | **2.95 (pooled)** | **chr1 in 47/47** |

Reads recruited against the rCRS alone (Pass 1) were realigned to the complete GRCh38 genome (Pass 2).

Relocated reads are primary alignments assigned to a nuclear chromosome, interpreted as NUMT-derived.

The pooled relocated fraction is 2.95% (115,302 of 3,908,080 reads); the mean of per-sample fractions is 3.22%

Principal chromosomal destination denotes the chromosome to which the **majority** of the reads relocate, which does **not necessarily mean all reads are relocated to that chromosome**

### **Supplementary Table S3. Kraken2 taxonomic partition of candidate read pairs (total on all samples)**

| **Taxonomic assignment** | **Read pairs** | **Percentage of total** | **Disposition** |
| --- | --- | --- | --- |
| Homo sapiens | 1,685,329 | 93.411 | Retained |
| Unclassified | 106,204 | 5.886 | Retained |
| Classified above domain level | 11,860 | 0.657 | Retained |
| Bacteria | 811 | 0.045 | Removed |
| Archaea | 1 | 0.000 | Removed |
| Viruses | 0 | 0 | Removed |
| Other Eukaryota | 0 | 0 | Retained |
| **TOTAL** | **1,804,205** | **100.000** |  |

Read pairs retained on chrM after competitive realignment were classified with Kraken2 v2.1.3 (Standard-8 database, build 2024-05-26).

Only pairs assigned to a taxon within Bacteria, Archaea or Viruses were removed; pairs classified as Homo sapiens, left unclassified, or classified only above domain level were retained.

### **Supplementary Table S4. Per-sample Kraken2 classification and removal**

| **Sample** | **Reads before classification** | **Reads after classification** | **Reads removed** | **Removed (%)** | **Bacterial reads** | **Eukaryota-assigned reads** |
| --- | --- | --- | --- | --- | --- | --- |
| 387 | 25,124 | 25,106 | 18 | 0.072 | 9 | 11,578 |
| 388 | 45,953 | 45,901 | 52 | 0.113 | 26 | 20,844 |
| 389 | 121,896 | 121,890 | 6 | 0.005 | 3 | 55,969 |
| 390 | 50,817 | 50,759 | 58 | 0.114 | 29 | 22,868 |
| 391 | 31,921 | 31,907 | 14 | 0.044 | 7 | 14,785 |
| 392 | 42,354 | 42,350 | 4 | 0.009 | 2 | 19,390 |
| 393 | 196,605 | 196,435 | 170 | 0.086 | 84 | 90,657 |
| 394 | 128,853 | 128,803 | 50 | 0.039 | 25 | 58,245 |
| 395 | 101,389 | 101,379 | 10 | 0.010 | 5 | 47,149 |
| 396 | 26,973 | 26,919 | 54 | 0.200 | 27 | 12,610 |
| 397 | 30,642 | 30,582 | 60 | 0.196 | 30 | 14,080 |
| 398 | 42,857 | 42,841 | 16 | 0.037 | 8 | 19,203 |
| 399 | 133,954 | 133,856 | 98 | 0.073 | 49 | 62,023 |
| 400 | 24,765 | 24,737 | 28 | 0.113 | 14 | 11,450 |
| 401 | 21,058 | 21,046 | 12 | 0.057 | 6 | 9,622 |
| 402 | 206,679 | 206,591 | 88 | 0.043 | 43 | 93,846 |
| 403 | 2,613 | 2,613 | 0 | 0.000 | 0 | 1,237 |
| 404 | 14,190 | 14,180 | 10 | 0.070 | 5 | 6,570 |
| 405 | 117,380 | 117,308 | 72 | 0.061 | 36 | 54,420 |
| 406 | 34,706 | 34,692 | 14 | 0.040 | 7 | 16,056 |
| 407 | 2,795 | 2,777 | 18 | 0.644 | 9 | 1,056 |
| 408 | 81,312 | 81,245 | 67 | 0.082 | 33 | 36,633 |
| 409 | 18,006 | 18,002 | 4 | 0.022 | 2 | 8,223 |
| 410 | 110,319 | 110,219 | 100 | 0.091 | 50 | 51,267 |
| 411 | 28,387 | 28,379 | 8 | 0.028 | 4 | 13,234 |
| 412 | 21,034 | 21,020 | 14 | 0.067 | 7 | 9,800 |
| 413 | 34,838 | 34,804 | 34 | 0.098 | 17 | 16,277 |
| 414 | 197,796 | 197,734 | 62 | 0.031 | 31 | 93,742 |
| 415 | 78,806 | 78,796 | 10 | 0.013 | 5 | 36,485 |
| 416 | 63,850 | 63,826 | 24 | 0.038 | 12 | 29,620 |
| 417 | 83,365 | 83,357 | 8 | 0.010 | 3 | 38,759 |
| 418 | 292,576 | 292,290 | 286 | 0.098 | 143 | 137,474 |
| 419 | 9,455 | 9,453 | 2 | 0.021 | 1 | 4,436 |
| 420 | 26,335 | 26,333 | 2 | 0.008 | 1 | 12,372 |
| 421 | 48,690 | 48,682 | 8 | 0.016 | 4 | 22,660 |
| 422 | 24,159 | 24,145 | 14 | 0.058 | 7 | 11,120 |
| 423 | 35,604 | 35,582 | 22 | 0.062 | 11 | 16,580 |
| 424 | 80,083 | 80,057 | 26 | 0.032 | 13 | 36,964 |
| 425 | 540,977 | 540,943 | 34 | 0.006 | 17 | 251,647 |
| 426 | 15,546 | 15,544 | 2 | 0.013 | 1 | 7,280 |
| 427 | 8,862 | 8,862 | 0 | 0.000 | 0 | 4,134 |
| 428 | 97,899 | 97,883 | 16 | 0.016 | 8 | 45,370 |
| 429 | 8,458 | 8,458 | 0 | 0.000 | 0 | 3,946 |
| 431 | 130,714 | 130,698 | 16 | 0.012 | 8 | 59,338 |
| 432 | 45,670 | 45,668 | 2 | 0.004 | 1 | 21,625 |
| 433 | 37,626 | 37,626 | 0 | 0.000 | 0 | 17,154 |
| 434 | 118,236 | 118,220 | 16 | 0.014 | 8 | 55,531 |

Reads removed are those assigned to Bacteria, Archaea or Viruses. Eukaryota-assigned reads, almost entirely Homo sapiens, were retained. Removal did not exceed 0.644% in any sample.

### **Supplementary Table S5. Pooled substitution-specific beta-binomial error model**

| **Substitution** | **Class** | **α** | **β** | **Mean error rate** | **Phred (Q)** | **Background observations** | **Ratio to global rate** |
| --- | --- | --- | --- | --- | --- | --- | --- |
| T>A | Transversion | 3.557 | 5,066 | 7.02e-04 | 31.5 | 165,268 | 3.54 |
| A>T | Transversion | 5.707 | 16,722 | 3.41e-04 | 34.7 | 212,861 | 1.72 |
| G>A | Transition | 2.086 | 8,489 | 2.46e-04 | 36.1 | 89,705 | 1.24 |
| T>C | Transition | 2.101 | 11,941 | 1.76e-04 | 37.5 | 171,708 | 0.89 |
| C>T | Transition | 2.534 | 15,625 | 1.62e-04 | 37.9 | 218,926 | 0.82 |
| A>G | Transition | 2.066 | 13,741 | 1.50e-04 | 38.2 | 215,090 | 0.76 |
| C>A | Transversion | 2.615 | 17,965 | 1.46e-04 | 38.4 | 219,572 | 0.73 |
| A>C | Transversion | 0.418 | 2,872 | 1.46e-04 | 38.4 | 215,001 | 0.73 |
| G>T | Transversion | 2.772 | 22,070 | 1.26e-04 | 39.0 | 90,729 | 0.63 |
| G>C | Transversion | 1.067 | 9,310 | 1.15e-04 | 39.4 | 90,830 | 0.58 |
| T>G | Transversion | 0.350 | 4,832 | 7.25e-05 | 41.4 | 172,936 | 0.37 |
| C>G | Transversion | 1.241 | 22,693 | 5.47e-05 | 42.6 | 220,608 | 0.28 |
| **Global (all classes)** | **—** | **0.774** | **3,902** | **1.98e-04** | **37.0** | **2,083,234** | **1.00** |

Background observations are non-reference alleles below 0.5% allele fraction at positions with depth ≥50, pooled across all 47 samples.

Each substitution class was fitted independently by maximum likelihood with the concentration parameter (α+β) bounded at 10⁶; all twelve classes exceeded the 500-observation minimum.

Rates span 12.8-fold, from T>A (Q31.5) to C>G (Q42.6).

### **Supplementary Table S6. Transition/transversion ratio and T>A calls across allele-fraction thresholds**

| **Minimum VAF (%)** | **Calls (n)** | **Transitions** | **Transversions** | **Ti/Tv** | **T>A calls** |
| --- | --- | --- | --- | --- | --- |
| ≥1 | 134 | 40 | 94 | 0.43 | 57 |
| ≥2 | 43 | 19 | 24 | 0.79 | 12 |
| ≥3 | 30 | 17 | 13 | 1.31 | 5 |
| ≥5 | 16 | 11 | 5 | 2.20 | 0 |
| ≥7 | 12 | 8 | 4 | 2.00 | 0 |
| ≥10 | 10 | 7 | 3 | 2.33 | 0 |

Computed from the 134 heteroplasmic calls.

The gnomAD v3.1 chrM Ti/Tv is 3.51. T>A calls fall to zero at 5%, where Ti/Tv reaches 2.20 and then plateaus; the maximum T>A allele fraction was 4.72%.

The 5% threshold was selected from these data, so this table describes the separation rather than an independent test.

### **Supplementary Table S7. All heteroplasmic calls with functional, population and lineage annotation (n = 134)**

| **Sample** | **Hg** | **Variant** | **VAF (%)** | **Depth** | **Alt reads** | **Fwd/Rev alt** | **P (error model)** | **Strand-bias P** | **Region** | **Gene** | **AA change** | **Effect** | **gnomAD AF hom** | **gnomAD AF het** | **gnomAD filter** | **Population status** | **Lineage position** | **Lineage verdict** |
| --- | --- | --- | --- | --- | --- | --- | --- | --- | --- | --- | --- | --- | --- | --- | --- | --- | --- | --- |
| 387 | R | m.5131T>A | 1.91 | 210 | 4 | 1/3 | 0.004 | 0.316 | Protein-coding | MT-ND2 | L221* | Nonsense | — | — | — | Novel | No | Private |
| 388 | R | m.13664T>A | 1.54 | 260 | 4 | 4/0 | 0.012 | 0.044 | Protein-coding | MT-ND5 | I443N | Missense | — | — | — | Novel | No | Private |
| 388 | R | m.13868T>A | 2.37 | 253 | 6 | 6/0 | 0.002 | 0.035 | Protein-coding | MT-ND5 | L511H | Missense | — | — | — | Novel | No | Private |
| 389 | M | m.9545A>G | 1.15 | 349 | 4 | 1/3 | 7.78e-04 | 0.484 | Protein-coding | MT-CO3 | G113G | Synonymous | 0.020740 | 0.000071 | PASS | Known polymorphism | No | Private |
| 389 | M | m.12310A>T | 1.44 | 901 | 13 | 11/2 | 1.83e-04 | 0.147 | tRNA | MT-TL2 | — | Non-coding | — | — | — | Novel | No | Private |
| 389 | M | m.12317T>G | 1.05 | 854 | 9 | 8/1 | 0.001 | 0.165 | tRNA | MT-TL2 | — | Non-coding | — | — | — | Novel | No | Private |
| 389 | M | m.16024T>A | 1.04 | 769 | 8 | 7/1 | 6.13e-06 | 0.147 | Control region | — | — | Non-coding | — | — | — | Novel | No | Private |
| 390 | M | m.877G>A | 1.57 | 254 | 4 | 2/2 | 0.018 | 1.000 | MT-RNR1 | MT-RNR1 | — | rRNA | 0.000000 | 0.000018 | PASS | Seen at heteroplasmy | No | Private |
| 390 | M | m.4183T>A | 1.66 | 241 | 4 | 3/1 | 2.00e-04 | 0.338 | Protein-coding | MT-ND1 | F293I | Missense | — | — | — | Novel | No | Private |
| 390 | M | m.6852G>A | 39.25 | 107 | 42 | 17/25 | 6.57e-113 | 0.694 | Protein-coding | MT-CO1 | G317S | Missense | 0.000248 | 0.000089 | PASS | Seen at heteroplasmy | No | Private |
| 390 | M | m.7549T>A | 4.72 | 212 | 10 | 9/1 | 2.31e-05 | 0.045 | tRNA | MT-TD | — | Non-coding | — | — | — | Novel | No | Private |
| 390 | M | m.12274A>T | 1.54 | 260 | 4 | 3/1 | 0.001 | 0.630 | tRNA | MT-TL2 | — | Non-coding | — | — | — | Novel | No | Private |
| 390 | M | m.12310A>T | 1.72 | 290 | 5 | 4/1 | 0.010 | 0.402 | tRNA | MT-TL2 | — | Non-coding | — | — | — | Novel | No | Private |
| 390 | M | m.12813C>T | 2.51 | 239 | 6 | 4/2 | 1.00e-09 | 0.406 | Protein-coding | MT-ND5 | Y159Y | Synonymous | 0.000673 | 0.000000 | PASS | Rare | No | Private |
| 391 | R | m.4939T>A | 1.70 | 236 | 4 | 4/0 | 8.59e-05 | 0.125 | Protein-coding | MT-ND2 | L157H | Missense | — | — | — | Novel | No | Private |
| 392 | M | m.3998T>A | 1.35 | 297 | 4 | 4/0 | 0.040 | 0.132 | Protein-coding | MT-ND1 | I231N | Missense | — | — | — | Novel | No | Private |
| 392 | M | m.13572T>C | 15.21 | 309 | 47 | 21/26 | <1e-300 | 0.527 | Protein-coding | MT-ND5 | T412T | Synonymous | 0.000709 | 0.000106 | PASS | Seen at heteroplasmy | No | Private |
| 392 | M | m.15530T>A | 1.13 | 353 | 4 | 4/0 | 0.011 | 0.033 | Protein-coding | MT-CYB | L262M | Missense | 0.000018 | 0.000000 | PASS | Rare | No | Shared, other macrohaplogroup only |
| 392 | M | m.15645T>A | 1.54 | 260 | 4 | 4/0 | 0.002 | 0.031 | Protein-coding | MT-CYB | I300N | Missense | 0.000000 | 0.000000 | npg | Rare | No | Private |
| 393 | R | m.3917A>G | 1.05 | 1139 | 12 | 9/3 | 9.61e-05 | 0.160 | Protein-coding | MT-ND1 | E204G | Missense | 0.000000 | 0.000000 | npg | Rare | No | Private |
| 393 | R | m.4833A>G | 1.38 | 727 | 10 | 5/5 | 1.44e-13 | 1.000 | Protein-coding | MT-ND2 | T122A | Missense | 0.001524 | 0.000000 | PASS | Known polymorphism | No | Private |
| 394 | M | m.316G>C | 1.51 | 331 | 5 | 1/4 | 0.021 | 0.477 | Control region | — | — | Non-coding | 0.000042 | 0.000021 | artifact_prone_site | gnomAD artifact-prone site | No | Private |
| 394 | M | m.864T>C | 1.26 | 556 | 7 | 2/5 | 0.027 | 0.269 | MT-RNR1 | MT-RNR1 | — | rRNA | 0.000000 | 0.000018 | PASS | Seen at heteroplasmy | No | Private |
| 394 | M | m.13602T>C | 42.57 | 855 | 364 | 180/184 | <1e-300 | 0.534 | Protein-coding | MT-ND5 | Y422Y | Synonymous | 0.000780 | 0.000035 | PASS | Seen at heteroplasmy | No | Private |
| 395 | R | m.523A>C | 2.92 | 137 | 4 | 1/3 | 1.59e-04 | 1.000 | Control region | — | — | Non-coding | 0.000000 | 0.000000 | npg | Rare | No | Private |
| 395 | R | m.1201A>G | 1.59 | 251 | 4 | 0/4 | 5.41e-07 | 0.158 | MT-RNR1 | MT-RNR1 | — | rRNA | 0.000000 | 0.000089 | PASS | Seen at heteroplasmy | No | Private |
| 395 | R | m.2339G>A | 2.15 | 790 | 17 | 9/8 | 2.25e-25 | 0.623 | MT-RNR2 | MT-RNR2 | — | rRNA | 0.000479 | 0.000035 | PASS | Seen at heteroplasmy | No | Private |
| 395 | R | m.3166T>A | 1.00 | 400 | 4 | 2/2 | 5.97e-04 | 1.000 | MT-RNR2 | MT-RNR2 | — | rRNA | — | — | — | Novel | No | Private |
| 396 | R | m.13844A>G | 3.12 | 128 | 4 | 2/2 | 0.013 | 1.000 | Protein-coding | MT-ND5 | D503G | Missense | 0.000000 | 0.000000 | npg | Rare | No | Private |
| 396 | R | m.15428G>C | 1.92 | 208 | 4 | 0/4 | 1.72e-04 | 0.055 | Protein-coding | MT-CYB | D228H | Missense | — | — | — | Novel | No | Private |
| 397 | R | m.9219T>A | 1.75 | 228 | 4 | 3/1 | 0.010 | 0.332 | Protein-coding | MT-CO3 | S5T | Missense | — | — | — | Novel | No | Private |
| 397 | R | m.15831T>A | 2.26 | 177 | 4 | 2/2 | 1.82e-04 | 1.000 | Protein-coding | MT-CYB | I362N | Missense | — | — | — | Novel | No | Private |
| 397 | R | m.16060G>A | 4.02 | 199 | 8 | 7/1 | 0.032 | 0.147 | Control region | — | — | Non-coding | — | — | — | Novel | No | Private |
| 398 | M | m.4078T>A | 1.19 | 336 | 4 | 4/0 | 0.010 | 0.039 | Protein-coding | MT-ND1 | Y258N | Missense | — | — | — | Novel | No | Private |
| 398 | M | m.6419A>C | 3.25 | 154 | 5 | 0/5 | 0.014 | 0.019 | Protein-coding | MT-CO1 | K172N | Missense | 0.000000 | 0.000000 | npg | Rare | No | Private |
| 398 | M | m.8143T>A | 3.88 | 103 | 4 | 1/3 | 0.015 | 0.067 | Protein-coding | MT-CO2 | A186A | Synonymous | — | — | — | Novel | No | Private |
| 398 | M | m.11088T>A | 1.79 | 280 | 5 | 4/1 | 2.60e-05 | 0.388 | Protein-coding | MT-ND4 | F110Y | Missense | 0.000000 | 0.000000 | npg | Rare | No | Private |
| 398 | M | m.14345T>A | 1.32 | 302 | 4 | 4/0 | 0.006 | 0.028 | Protein-coding | MT-ND6 | D66V | Missense | — | — | — | Novel | No | Private |
| 398 | M | m.16024T>A | 2.54 | 275 | 7 | 3/4 | 5.74e-08 | 1.000 | Control region | — | — | Non-coding | — | — | — | Novel | No | Private |
| 399 | M | m.22T>A | 1.27 | 316 | 4 | 4/0 | 0.001 | 0.308 | Control region | — | — | Non-coding | — | — | — | Novel | No | Private |
| 399 | M | m.316G>C | 1.40 | 429 | 6 | 0/6 | 0.016 | 1.000 | Control region | — | — | Non-coding | 0.000042 | 0.000021 | artifact_prone_site | gnomAD artifact-prone site | No | Private |
| 399 | M | m.7549T>A | 1.96 | 562 | 11 | 9/2 | 0.014 | 0.223 | tRNA | MT-TD | — | Non-coding | — | — | — | Novel | No | Private |
| 399 | M | m.12310A>T | 1.18 | 851 | 10 | 2/8 | 0.005 | 0.024 | tRNA | MT-TL2 | — | Non-coding | — | — | — | Novel | No | Private |
| 400 | M | m.16060G>A | 4.07 | 172 | 7 | 6/1 | 0.041 | 0.124 | Control region | — | — | Non-coding | — | — | — | Novel | No | Private |
| 400 | M | m.16093T>C | 90.81 | 185 | 168 | 97/71 | 9.06e-293 | 1.000 | Control region | — | — | Non-coding | 0.053108 | 0.011641 | PASS | Known polymorphism | Yes | Shared, both macrohaplogroups |
| 402 | R | m.546A>C | 1.03 | 483 | 5 | 4/1 | 0.002 | 0.033 | Control region | — | — | Non-coding | 0.000000 | 0.000000 | npg | Rare | No | Private |
| 402 | R | m.8121T>A | 1.99 | 503 | 10 | 4/6 | 0.007 | 0.182 | Protein-coding | MT-CO2 | L179Q | Missense | — | — | — | Novel | No | Private |
| 405 | R | m.10577A>G | 1.82 | 603 | 11 | 6/5 | <1e-300 | 1.000 | Protein-coding | MT-ND4L | M36M | Synonymous | 0.000018 | 0.000018 | PASS | Seen at heteroplasmy | No | Private |
| 405 | R | m.15196A>G | 1.24 | 482 | 6 | 4/2 | 0.004 | 0.688 | Protein-coding | MT-CYB | L150L | Synonymous | 0.000159 | 0.000018 | PASS | Seen at heteroplasmy | No | Private |
| 408 | R | m.13868T>A | 1.26 | 476 | 6 | 5/1 | 0.043 | 0.226 | Protein-coding | MT-ND5 | L511H | Missense | — | — | — | Novel | No | Private |
| 409 | R | m.12310A>T | 5.26 | 95 | 5 | 4/1 | 7.25e-05 | 1.000 | tRNA | MT-TL2 | — | Non-coding | — | — | — | Novel | No | Private |
| 409 | R | m.13844A>G | 3.97 | 151 | 6 | 6/0 | 6.92e-04 | 0.010 | Protein-coding | MT-ND5 | D503G | Missense | 0.000000 | 0.000000 | npg | Rare | No | Private |
| 410 | M | m.3523A>C | 1.04 | 385 | 4 | 3/1 | 0.002 | 0.138 | Protein-coding | MT-ND1 | T73P | Missense | 0.000000 | 0.000000 | npg | Rare | No | Private |
| 410 | M | m.4003A>G | 1.89 | 792 | 15 | 7/8 | <1e-300 | 1.000 | Protein-coding | MT-ND1 | M233V | Missense | — | — | — | Novel | No | Private |
| 410 | M | m.9523T>A | 1.02 | 393 | 4 | 3/1 | 0.022 | 0.131 | Protein-coding | MT-CO3 | L106Q | Missense | — | — | — | Novel | No | Private |
| 410 | M | m.10143G>A | 4.16 | 769 | 32 | 12/20 | 5.33e-51 | 0.149 | Protein-coding | MT-ND3 | G29S | Missense | 0.004059 | 0.000071 | PASS | Known polymorphism | No | Private |
| 410 | M | m.12974T>A | 1.00 | 398 | 4 | 4/0 | 0.034 | 0.041 | Protein-coding | MT-ND5 | L213Q | Missense | — | — | — | Novel | No | Private |
| 410 | M | m.13868T>A | 1.16 | 605 | 7 | 7/0 | 0.043 | 0.046 | Protein-coding | MT-ND5 | L511H | Missense | — | — | — | Novel | No | Private |
| 411 | M | m.3998T>A | 1.70 | 236 | 4 | 4/0 | 0.019 | 0.136 | Protein-coding | MT-ND1 | I231N | Missense | — | — | — | Novel | No | Private |
| 411 | M | m.10237T>C | 6.52 | 138 | 9 | 3/6 | <1e-300 | 1.000 | Protein-coding | MT-ND3 | I60T | Missense | 0.001790 | 0.000160 | PASS | Known polymorphism | No | Private |
| 411 | M | m.11616T>A | 2.53 | 158 | 4 | 3/1 | 1.86e-05 | 0.644 | Protein-coding | MT-ND4 | I286N | Missense | — | — | — | Novel | No | Private |
| 411 | M | m.14173T>A | 1.40 | 356 | 5 | 4/1 | 2.85e-04 | 0.201 | Protein-coding | MT-ND6 | V9V | Synonymous | — | — | — | Novel | No | Private |
| 411 | M | m.15560T>A | 1.52 | 263 | 4 | 4/0 | 0.005 | 0.023 | Protein-coding | MT-CYB | W272* | Nonsense | — | — | — | Novel | No | Private |
| 412 | M | m.8074A>G | 5.13 | 78 | 4 | 1/3 | 6.44e-09 | 0.341 | Protein-coding | MT-CO2 | W163W | Synonymous | 0.000195 | 0.000035 | PASS | Seen at heteroplasmy | No | Private |
| 412 | M | m.16060G>A | 3.88 | 206 | 8 | 8/0 | 0.038 | 0.022 | Control region | — | — | Non-coding | — | — | — | Novel | No | Private |
| 413 | M | m.246T>C | 23.64 | 165 | 39 | 24/15 | 3.78e-95 | 0.142 | Control region | — | — | Non-coding | 0.001453 | 0.000106 | PASS | Known polymorphism | No | Private |
| 413 | M | m.13169T>A | 1.70 | 235 | 4 | 3/1 | 0.002 | 1.000 | Protein-coding | MT-ND5 | L278Q | Missense | — | — | — | Novel | No | Private |
| 413 | M | m.14469A>C | 2.42 | 207 | 5 | 2/3 | 9.59e-07 | 1.000 | Protein-coding | MT-ND6 | Y107D | Missense | — | — | — | Novel | No | Private |
| 414 | M | m.8121T>A | 1.56 | 577 | 9 | 3/6 | 0.047 | 0.021 | Protein-coding | MT-CO2 | L179Q | Missense | — | — | — | Novel | No | Private |
| 415 | M | m.4842A>G | 1.51 | 332 | 5 | 3/2 | 0.047 | 1.000 | Protein-coding | MT-ND2 | T125A | Missense | 0.000018 | 0.000000 | PASS | Rare | No | Private |
| 415 | M | m.6432A>C | 2.31 | 346 | 8 | 1/7 | 0.006 | 0.023 | Protein-coding | MT-CO1 | T177P | Missense | — | — | — | Novel | No | Private |
| 415 | M | m.7789G>A | 47.72 | 570 | 272 | 113/159 | <1e-300 | 0.446 | Protein-coding | MT-CO2 | L68L | Synonymous | 0.010988 | 0.000018 | PASS | Known polymorphism | No | Private |
| 415 | M | m.8134T>A | 3.03 | 165 | 5 | 5/0 | 0.020 | 0.324 | Protein-coding | MT-CO2 | T183T | Synonymous | — | — | — | Novel | No | Private |
| 415 | M | m.11464C>A | 1.08 | 461 | 5 | 3/2 | 0.010 | 1.000 | Protein-coding | MT-ND4 | L235L | Synonymous | 0.000018 | 0.000000 | PASS | Rare | No | Private |
| 415 | M | m.12317T>G | 1.42 | 562 | 8 | 8/0 | 3.65e-04 | 0.024 | tRNA | MT-TL2 | — | Non-coding | — | — | — | Novel | No | Private |
| 415 | M | m.12924A>C | 1.01 | 394 | 4 | 3/1 | 0.010 | 0.337 | Protein-coding | MT-ND5 | W196C | Missense | 0.000000 | 0.000000 | npg | Rare | No | Private |
| 415 | M | m.13133T>A | 1.09 | 367 | 4 | 3/1 | 0.001 | 0.634 | Protein-coding | MT-ND5 | L266Q | Missense | — | — | — | Novel | No | Private |
| 415 | M | m.13143T>A | 1.16 | 433 | 5 | 4/1 | 0.001 | 0.389 | Protein-coding | MT-ND5 | N269K | Missense | — | — | — | Novel | No | Private |
| 415 | M | m.15923A>G | 1.16 | 519 | 6 | 3/3 | 1.69e-08 | 1.000 | tRNA | MT-TT | — | Non-coding | 0.000000 | 0.000000 | npg | Rare | No | Private |
| 416 | R | m.864T>C | 1.82 | 274 | 5 | 2/3 | 0.015 | 0.672 | MT-RNR1 | MT-RNR1 | — | rRNA | 0.000000 | 0.000018 | PASS | Seen at heteroplasmy | No | Private |
| 416 | R | m.8315A>T | 1.16 | 344 | 4 | 4/0 | 0.037 | 0.126 | tRNA | MT-TK | — | Non-coding | — | — | — | Novel | No | Private |
| 417 | N | m.864T>C | 1.49 | 336 | 5 | 0/5 | 0.033 | 0.016 | MT-RNR1 | MT-RNR1 | — | rRNA | 0.000000 | 0.000018 | PASS | Seen at heteroplasmy | No | Private |
| 417 | N | m.6438T>A | 2.93 | 375 | 11 | 7/4 | 0.005 | 1.000 | Protein-coding | MT-CO1 | Y179N | Missense | — | — | — | Novel | No | Private |
| 417 | N | m.7943T>A | 1.10 | 364 | 4 | 2/2 | 0.015 | 1.000 | Protein-coding | MT-CO2 | S120T | Missense | — | — | — | Novel | No | Private |
| 417 | N | m.8143T>A | 2.66 | 188 | 5 | 3/2 | 0.030 | 0.161 | Protein-coding | MT-CO2 | A186A | Synonymous | — | — | — | Novel | No | Private |
| 417 | N | m.8303A>T | 1.16 | 344 | 4 | 3/1 | 0.003 | 0.629 | tRNA | MT-TK | — | Non-coding | — | — | — | Novel | No | Private |
| 417 | N | m.10458C>A | 1.21 | 496 | 6 | 1/5 | 6.59e-04 | 0.119 | tRNA | MT-TR | — | Non-coding | — | — | — | Novel | No | Private |
| 417 | N | m.13046T>A | 2.39 | 209 | 5 | 4/1 | 0.014 | 0.060 | Protein-coding | MT-ND5 | M237K | Missense | — | — | — | Novel | No | Private |
| 417 | N | m.15388T>A | 1.30 | 540 | 7 | 6/1 | 0.003 | 0.123 | Protein-coding | MT-CYB | H214Q | Missense | — | — | — | Novel | No | Private |
| 418 | R | m.292T>A | 4.30 | 1022 | 44 | 20/24 | 6.22e-30 | 0.046 | Control region | — | — | Non-coding | 0.000408 | 0.000000 | PASS | Rare | No | Private |
| 418 | R | m.3106C>A | 40.38 | 52 | 21 | 13/8 | <1e-300 | 0.773 | MT-RNR2 | MT-RNR2 | — | rRNA | — | — | — | Novel | No | Private |
| 419 | M | m.12127G>C | 30.00 | 60 | 18 | 8/10 | <1e-300 | 0.783 | Protein-coding | MT-ND4 | G456G | Synonymous | — | — | — | Novel | No | Private |
| 420 | R | m.5441A>C | 1.20 | 334 | 4 | 4/0 | 0.018 | 0.010 | Protein-coding | MT-ND2 | P324P | Synonymous | 0.000018 | 0.000000 | PASS | Rare | No | Private |
| 420 | R | m.8634T>A | 1.20 | 333 | 4 | 2/2 | 0.003 | 1.000 | Protein-coding | MT-ATP6 | Y36* | Nonsense | — | — | — | Novel | No | Private |
| 420 | R | m.8668T>A | 1.24 | 322 | 4 | 4/0 | 0.043 | 0.125 | Protein-coding | MT-ATP6 | W48* | Nonsense | — | — | — | Novel | No | Private |
| 420 | R | m.13651A>C | 2.10 | 190 | 4 | 4/0 | 0.038 | 0.030 | Protein-coding | MT-ND5 | T439P | Missense | 0.000053 | 0.000000 | PASS | Rare | No | Private |
| 420 | R | m.13781T>A | 1.59 | 251 | 4 | 4/0 | 6.46e-04 | 0.034 | Protein-coding | MT-ND5 | I482N | Missense | 0.000000 | 0.000000 | npg | Rare | No | Private |
| 420 | R | m.13869T>A | 1.47 | 272 | 4 | 4/0 | 0.010 | 0.152 | Protein-coding | MT-ND5 | L511L | Synonymous | — | — | — | Novel | No | Private |
| 420 | R | m.13874T>A | 1.68 | 298 | 5 | 2/3 | 0.001 | 0.653 | Protein-coding | MT-ND5 | M513K | Missense | — | — | — | Novel | No | Private |
| 420 | R | m.16060G>A | 7.18 | 209 | 15 | 12/3 | 8.08e-06 | 0.177 | Control region | — | — | Non-coding | — | — | — | Novel | No | Private |
| 421 | M | m.10081T>A | 1.61 | 248 | 4 | 3/1 | 1.04e-04 | 1.000 | Protein-coding | MT-ND3 | M8K | Missense | — | — | — | Novel | No | Private |
| 421 | M | m.10323A>G | 1.81 | 221 | 4 | 4/0 | 2.94e-05 | 0.048 | Protein-coding | MT-ND3 | M89V | Missense | 0.000018 | 0.000000 | PASS | Rare | No | Private |
| 421 | M | m.15477T>A | 1.12 | 446 | 5 | 5/0 | 0.002 | 0.030 | Protein-coding | MT-CYB | L244Q | Missense | — | — | — | Novel | No | Private |
| 421 | M | m.15560T>A | 1.06 | 377 | 4 | 4/0 | 0.017 | 0.025 | Protein-coding | MT-CYB | W272* | Nonsense | — | — | — | Novel | No | Private |
| 421 | M | m.16044T>C | 1.18 | 338 | 4 | 1/3 | 0.015 | 0.324 | Control region | — | — | Non-coding | 0.000000 | 0.000018 | PASS | Seen at heteroplasmy | No | Private |
| 423 | R | m.3892A>C | 1.84 | 218 | 4 | 0/4 | 5.37e-04 | 0.122 | Protein-coding | MT-ND1 | T196P | Missense | 0.000000 | 0.000000 | npg | Rare | No | Private |
| 423 | R | m.5131T>A | 1.44 | 278 | 4 | 1/3 | 0.010 | 0.369 | Protein-coding | MT-ND2 | L221* | Nonsense | — | — | — | Novel | No | Private |
| 423 | R | m.11529T>A | 1.82 | 220 | 4 | 4/0 | 5.20e-04 | 0.142 | Protein-coding | MT-ND4 | M257K | Missense | — | — | — | Novel | No | Private |
| 423 | R | m.11678T>A | 1.60 | 250 | 4 | 3/1 | 3.07e-04 | 0.627 | Protein-coding | MT-ND4 | W307* | Nonsense | — | — | — | Novel | No | Private |
| 423 | R | m.14325T>C | 1.53 | 261 | 4 | 2/2 | 1.21e-06 | 1.000 | Protein-coding | MT-ND6 | N59D | Missense | 0.001382 | 0.000018 | PASS | Known polymorphism | No | Private |
| 424 | M | m.251G>A | 1.05 | 380 | 4 | 3/1 | 3.93e-05 | 0.365 | Control region | — | — | Non-coding | 0.000284 | 0.000160 | PASS | Seen at heteroplasmy | No | Private |
| 424 | M | m.15388T>A | 1.14 | 615 | 7 | 6/1 | 0.006 | 0.133 | Protein-coding | MT-CYB | H214Q | Missense | — | — | — | Novel | No | Private |
| 425 | M | m.297A>C | 9.16 | 393 | 36 | 7/29 | 7.39e-13 | 0.182 | Control region | — | — | Non-coding | 0.000000 | 0.000000 | npg | Rare | No | Private |
| 425 | M | m.300A>C | 3.38 | 384 | 13 | 4/9 | 8.66e-12 | 0.305 | Control region | — | — | Non-coding | 0.000000 | 0.000000 | npg | Rare | No | Private |
| 426 | M | m.10060T>G | 4.08 | 98 | 4 | 2/2 | 4.12e-06 | 1.000 | Protein-coding | MT-ND3 | M1* | Nonsense | — | — | — | Novel | No | Private |
| 427 | R | m.16060G>A | 6.35 | 63 | 4 | 4/0 | 0.029 | 0.288 | Control region | — | — | Non-coding | — | — | — | Novel | No | Private |
| 428 | M | m.6654T>G | 1.13 | 353 | 4 | 4/0 | 0.005 | 0.023 | Protein-coding | MT-CO1 | F251V | Missense | — | — | — | Novel | No | Private |
| 428 | M | m.7972A>C | 1.38 | 363 | 5 | 0/5 | 0.031 | 0.070 | Protein-coding | MT-CO2 | E129D | Missense | — | — | — | Novel | No | Private |
| 428 | M | m.8084A>C | 1.22 | 329 | 4 | 4/0 | 0.002 | 0.144 | Protein-coding | MT-CO2 | T167P | Missense | 0.000000 | 0.000018 | PASS | Seen at heteroplasmy | No | Lineage, own macrohaplogroup |
| 428 | M | m.12164G>A | 1.65 | 668 | 11 | 4/7 | <1e-300 | 0.764 | tRNA | MT-TH | — | Non-coding | 0.000230 | 0.000106 | PASS | Seen at heteroplasmy | No | Private |
| 428 | M | m.16487A>T | 1.11 | 359 | 4 | 0/4 | 3.56e-04 | 1.000 | Control region | — | — | Non-coding | — | — | — | Novel | No | Private |
| 431 | R | m.7976G>A | 1.41 | 425 | 6 | 2/4 | 8.69e-08 | 0.694 | Protein-coding | MT-CO2 | G131S | Missense | 0.000195 | 0.000106 | PASS | Seen at heteroplasmy | No | Private |
| 432 | M | m.3482A>C | 1.38 | 289 | 4 | 2/2 | 2.24e-04 | 1.000 | Protein-coding | MT-ND1 | E59A | Missense | — | — | — | Novel | No | Private |
| 432 | M | m.6438T>A | 3.06 | 196 | 6 | 3/3 | 0.030 | 0.675 | Protein-coding | MT-CO1 | Y179N | Missense | — | — | — | Novel | No | Private |
| 432 | M | m.9375T>A | 1.36 | 293 | 4 | 3/1 | 0.002 | 0.312 | Protein-coding | MT-CO3 | W57* | Nonsense | — | — | — | Novel | No | Private |
| 432 | M | m.10761T>A | 1.05 | 379 | 4 | 3/1 | 0.005 | 0.628 | Protein-coding | MT-ND4L | C98S | Missense | — | — | — | Novel | No | Private |
| 432 | M | m.14871T>A | 1.22 | 329 | 4 | 2/2 | 2.94e-04 | 1.000 | Protein-coding | MT-CYB | I42N | Missense | — | — | — | Novel | No | Private |
| 433 | M | m.12310A>T | 1.71 | 234 | 4 | 3/1 | 0.022 | 1.000 | tRNA | MT-TL2 | — | Non-coding | — | — | — | Novel | No | Private |
| 433 | M | m.15366A>G | 1.50 | 267 | 4 | 2/2 | 6.90e-07 | 1.000 | Protein-coding | MT-CYB | N207S | Missense | 0.000000 | 0.000000 | npg | Rare | No | Private |
| 433 | M | m.16183A>C | 10.77 | 65 | 7 | 0/7 | 1.33e-11 | 1.000 | Control region | — | — | Non-coding | 0.004697 | 0.100824 | PASS | Known polymorphism | Yes | Private |
| 434 | M | m.204T>C | 20.06 | 673 | 135 | 70/65 | 1.16e-04 | 0.119 | Control region | — | — | Non-coding | 0.066087 | 0.004985 | PASS | Known polymorphism | Yes | Shared, both macrohaplogroups |
| 434 | M | m.316G>C | 1.47 | 407 | 6 | 0/6 | 0.013 | 1.000 | Control region | — | — | Non-coding | 0.000042 | 0.000021 | artifact_prone_site | gnomAD artifact-prone site | No | Private |
| 434 | M | m.8298T>A | 1.16 | 518 | 6 | 5/1 | 0.002 | 0.096 | tRNA | MT-TK | — | Non-coding | — | — | — | Novel | No | Private |
| 434 | M | m.13787T>C | 1.03 | 389 | 4 | 2/2 | 5.39e-06 | 0.586 | Protein-coding | MT-ND5 | L484P | Missense | 0.000035 | 0.000053 | PASS | Seen at heteroplasmy | No | Private |

Hg, macrohaplogroup. VAF, variant allele fraction.

P (error model) is the upper-tail probability of the alternate count under the substitution-specific beta-binomial model; where the leave-one-out site background rate exceeded the substitution rate, the larger of the two tail probabilities was used (calls required P < 0.05).

Strand-bias P is from Fisher's exact test (calls with P ≤ 0.01 were rejected).

Protein-coding annotation uses the vertebrate mitochondrial genetic code (translation table 2).

Population status, assigned in order of precedence from the gnomAD v3.1 chrM table: gnomAD artifact-prone site (filter flag); known polymorphism, homoplasmic AF ≥ 0.001; seen at heteroplasmy, heteroplasmic AF > 0; rare, present in gnomAD at neither level; novel, absent from gnomAD.

Lineage position marks the 165 PhyloTree or homoplasmic positions.

Sixteen calls exceed the 5% detection limit and ten exceed gnomAD's 10% threshold.

**Note:** Region was reassigned from the pipeline output for 15 calls that lie within tRNA genes inside the protein-coding span (m.3307–15887): MT-TL2 (m.12274, m.12310, m.12317), MT-TD (m.7549), MT-TK (m.8298, m.8303, m.8315), MT-TR (m.10458) and MT-TH (m.12164).

### **Supplementary Table S8. Positions called heteroplasmic in more than one sample (n = 16)**

| **Variant** | **Class** | **Samples (n)** | **Sample IDs** | **Individual VAFs (%)** | **Median VAF (%)** | **Maximum VAF (%)** | **Macrohaplogroups of carriers** | **gnomAD status** |
| --- | --- | --- | --- | --- | --- | --- | --- | --- |
| m.12310A>T | Transversion | 5 | 389, 390, 399, 409, 433 | 5.26, 1.72, 1.71, 1.44, 1.18 | 1.71 | 5.26 | M 4, R 1 | Novel |
| m.16060G>A | Transition | 5 | 397, 400, 412, 420, 427 | 7.18, 6.35, 4.07, 4.02, 3.88 | 4.07 | 7.18 | M 2, R 3 | Novel |
| m.316G>C | Transversion | 3 | 394, 399, 434 | 1.51, 1.47, 1.40 | 1.47 | 1.51 | M 3 | gnomAD artifact-prone site |
| m.864T>C | Transition | 3 | 394, 416, 417 | 1.82, 1.49, 1.26 | 1.49 | 1.82 | M 1, N 1, R 1 | Seen at heteroplasmy |
| m.13868T>A | Transversion | 3 | 388, 408, 410 | 2.37, 1.26, 1.16 | 1.26 | 2.37 | M 1, R 2 | Novel |
| m.3998T>A | Transversion | 2 | 392, 411 | 1.70, 1.35 | 1.52 | 1.70 | M 2 | Novel |
| m.5131T>A | Transversion | 2 | 387, 423 | 1.91, 1.44 | 1.67 | 1.91 | R 2 | Novel |
| m.6438T>A | Transversion | 2 | 417, 432 | 3.06, 2.93 | 3.00 | 3.06 | M 1, N 1 | Novel |
| m.7549T>A | Transversion | 2 | 390, 399 | 4.72, 1.96 | 3.34 | 4.72 | M 2 | Novel |
| m.8121T>A | Transversion | 2 | 402, 414 | 1.99, 1.56 | 1.77 | 1.99 | M 1, R 1 | Novel |
| m.8143T>A | Transversion | 2 | 398, 417 | 3.88, 2.66 | 3.27 | 3.88 | M 1, N 1 | Novel |
| m.12317T>G | Transversion | 2 | 389, 415 | 1.42, 1.05 | 1.24 | 1.42 | M 2 | Novel |
| m.13844A>G | Transition | 2 | 396, 409 | 3.97, 3.12 | 3.55 | 3.97 | R 2 | Rare |
| m.15388T>A | Transversion | 2 | 417, 424 | 1.30, 1.14 | 1.22 | 1.30 | M 1, N 1 | Novel |
| m.15560T>A | Transversion | 2 | 411, 421 | 1.52, 1.06 | 1.29 | 1.52 | M 2 | Novel |
| m.16024T>A | Transversion | 2 | 389, 398 | 2.54, 1.04 | 1.79 | 2.54 | M 2 | Novel |

Thirteen of the 16 recurrent positions are transversions, ten of them T>A. No recurrent position has a median VAF above 4.07%.

Two positions have a call above the 5% limit: m.16060G>A (5 samples, 3.88–7.18%) and m.12310A>T (5 samples, 1.18–5.26%); neither is present in gnomAD v3.1 or homoplasmic in any sample, and carriers of each include both M and R macrohaplogroups.

The remaining 93 positions were private to one sample.

### **Supplementary Table S9. Heteroplasmic calls at lineage positions or shared with homoplasmic carriers in the samples**

| **Sample** | **Hg** | **Variant** | **VAF (%)** | **Depth** | **Region** | **Lineage position** | **Homoplasmic carriers (n)** | **Carriers, same Hg** | **Carriers, other Hg** | **Carrier haplogroups** | **Verdict** |
| --- | --- | --- | --- | --- | --- | --- | --- | --- | --- | --- | --- |
| 392 | M | m.15530T>A | 1.13 | 353 | Protein-coding | No | 1 | 0 | 1 | R | Shared, other macrohaplogroup only |
| 400 | M | m.16093T>C | 90.81 | 185 | Control region | Yes | 2 | 1 | 1 | M, R | Shared, both macrohaplogroups |
| 428 | M | m.8084A>C | 1.22 | 329 | Protein-coding | No | 1 | 1 | 0 | M | Lineage, own macrohaplogroup |
| 433 | M | m.16183A>C | 10.77 | 65 | Control region | Yes | 0 | 0 | 0 | — | Private |
| 434 | M | m.204T>C | 20.06 | 673 | Control region | Yes | 4 | 3 | 1 | M, M, M, N | Shared, both macrohaplogroups |

Each of the 134 calls was tabulated against the homoplasmic variants (VAF ≥95%) of all samples.

Lineage positions are 165 positions that are PhyloTree markers or homoplasmic in ≥2 samples; calls there were flagged, not removed.

Across all 134 calls: Private, 130; Shared, both macrohaplogroups, 2; Lineage, own macrohaplogroup, 1; Shared, other macrohaplogroup only, 1. Only the calls not classified as private, or falling at a lineage position, are listed; the full classification is given in Table S7.

### **Supplementary Table S10. Distribution of heteroplasmic calls by genomic region**

| **Region** | **Calls (n)** | **Samples (n)** | **Median VAF (%)** | **Calls ≥5% VAF (n)** |
| --- | --- | --- | --- | --- |
| Protein-coding | 87 | 33 | 1.56 | 7 |
| Control region | 23 | 19 | 3.38 | 7 |
| MT-RNR1 | 5 | 5 | 1.57 | 0 |
| MT-RNR2 | 3 | 2 | 2.15 | 1 |
| tRNA | 16 | 10 | 1.43 | 1 |
| **Total** | **134** | **40** | **1.58** | **16** |

Regions: control region m.16024–576; MT-RNR1 m.648-1601; MT-RNR2 m.1671-3229; protein-coding genes within m.3307-15887; tRNA genes by rCRS coordinates.

Protein-coding calls comprised 62 missense, 16 synonymous and 9 nonsense changes.

**Note:** Counts differ from the pipeline region summary (protein-coding: 102, tRNA: 1), which assigned the 15 tRNA calls within m.3307-15887 to the protein-coding region; see Table S7.

### **Supplementary Table S11. Macrohaplogroup assignment**

| **Sample** | **Macrohaplogroup** | **Markers typed** | **Non-rCRS / typed** | **Basis of assignment** |
| --- | --- | --- | --- | --- |
| 387 | R | 11/11 | 0/11 | Inside N (5/5 rCRS); R by m.12705 |
| 388 | R | 11/11 | 0/11 | Inside N (5/5 rCRS); R by m.12705 |
| 389 | M | 11/11 | 11/11 | Outside N (5/5 non-rCRS); M markers 4/4 derived |
| 390 | M | 11/11 | 11/11 | Outside N (5/5 non-rCRS); M markers 4/4 derived |
| 391 | R | 11/11 | 0/11 | Inside N (5/5 rCRS); R by m.12705 |
| 392 | M | 11/11 | 11/11 | Outside N (5/5 non-rCRS); M markers 4/4 derived |
| 393 | R | 11/11 | 0/11 | Inside N (5/5 rCRS); R by m.12705 |
| 394 | M | 11/11 | 11/11 | Outside N (5/5 non-rCRS); M markers 4/4 derived |
| 395 | R | 11/11 | 0/11 | Inside N (5/5 rCRS); R by m.12705 |
| 396 | R | 11/11 | 0/11 | Inside N (5/5 rCRS); R by m.12705 |
| 397 | R | 11/11 | 0/11 | Inside N (5/5 rCRS); R by m.12705 |
| 398 | M | 11/11 | 11/11 | Outside N (5/5 non-rCRS); M markers 4/4 derived |
| 399 | M | 11/11 | 10/11 | Outside N (5/5 non-rCRS); M markers 4/4 derived |
| 400 | M | 11/11 | 11/11 | Outside N (5/5 non-rCRS); M markers 4/4 derived |
| 401 | R | 11/11 | 0/11 | Inside N (5/5 rCRS); R by m.12705 |
| 402 | R | 11/11 | 1/11 | Inside N (4/5 rCRS); R by m.12705 |
| 403 | R | 7/11 | 0/7 | Inside N (4/4 rCRS); R by m.12705 |
| 404 | R | 11/11 | 0/11 | Inside N (5/5 rCRS); R by m.12705 |
| 405 | R | 11/11 | 0/11 | Inside N (5/5 rCRS); R by m.12705 |
| 406 | R | 11/11 | 0/11 | Inside N (5/5 rCRS); R by m.12705 |
| 407 | Unassigned | 6/11 | 6/6 | N block typed at 2/5 (<3) |
| 408 | R | 11/11 | 0/11 | Inside N (5/5 rCRS); R by m.12705 |
| 409 | R | 11/11 | 0/11 | Inside N (5/5 rCRS); R by m.12705 |
| 410 | M | 11/11 | 11/11 | Outside N (5/5 non-rCRS); M markers 4/4 derived |
| 411 | M | 11/11 | 11/11 | Outside N (5/5 non-rCRS); M markers 4/4 derived |
| 412 | M | 11/11 | 11/11 | Outside N (5/5 non-rCRS); M markers 4/4 derived |
| 413 | M | 11/11 | 11/11 | Outside N (5/5 non-rCRS); M markers 4/4 derived |
| 414 | M | 11/11 | 11/11 | Outside N (5/5 non-rCRS); M markers 4/4 derived |
| 415 | M | 11/11 | 11/11 | Outside N (5/5 non-rCRS); M markers 4/4 derived |
| 416 | R | 11/11 | 0/11 | Inside N (5/5 rCRS); R by m.12705 |
| 417 | N (non-R) | 11/11 | 2/11 | Inside N (5/5 rCRS); non-R by m.12705 |
| 418 | R | 11/11 | 0/11 | Inside N (5/5 rCRS); R by m.12705 |
| 419 | M | 11/11 | 11/11 | Outside N (5/5 non-rCRS); M markers 4/4 derived |
| 420 | R | 11/11 | 0/11 | Inside N (5/5 rCRS); R by m.12705 |
| 421 | M | 11/11 | 11/11 | Outside N (5/5 non-rCRS); M markers 4/4 derived |
| 422 | R | 11/11 | 0/11 | Inside N (5/5 rCRS); R by m.12705 |
| 423 | R | 11/11 | 0/11 | Inside N (5/5 rCRS); R by m.12705 |
| 424 | M | 11/11 | 11/11 | Outside N (5/5 non-rCRS); M markers 4/4 derived |
| 425 | M | 11/11 | 11/11 | Outside N (5/5 non-rCRS); M markers 4/4 derived |
| 426 | M | 11/11 | 11/11 | Outside N (5/5 non-rCRS); M markers 4/4 derived |
| 427 | R | 11/11 | 0/11 | Inside N (5/5 rCRS); R by m.12705 |
| 428 | M | 11/11 | 11/11 | Outside N (5/5 non-rCRS); M markers 4/4 derived |
| 429 | R | 11/11 | 0/11 | Inside N (5/5 rCRS); R by m.12705 |
| 431 | R | 11/11 | 1/11 | Inside N (4/5 rCRS); R by m.12705 |
| 432 | M | 11/11 | 11/11 | Outside N (5/5 non-rCRS); M markers 4/4 derived |
| 433 | M | 11/11 | 11/11 | Outside N (5/5 non-rCRS); M markers 4/4 derived |
| 434 | M | 11/11 | 11/11 | Outside N (5/5 non-rCRS); M markers 4/4 derived |

Assignment used eleven diagnostic rCRS positions: five separating N from L3/M (m.8701, 9540, 10398, 10873, 15301), four defining M (m.489, 10400, 14783, 15043) and two separating R from other N lineages (m.12705 primary, m.16223 support).

A marker was typed when ≥10 reads passed base quality ≥20 and mapping quality ≥30 and the major allele made up ≥80% of them.

A sample was assigned only when ≥3 of the 5 N-block markers were typed and a majority pointed one way; outside N, ≥2 derived M markers were required for M.

The rCRS belongs to haplogroup H (within R), so R samples carry reference alleles at the diagnostic positions.

Allele-level detail is given in Table S12. Distribution: M 22, R 23, N (non-R) 1, unassigned 1.

**Note:** Samples 403 and 407 fell below the 50× calling threshold. Sample 403 was typed at 7 of 11 markers and assigned R on four concordant N-block markers. Sample 407 was typed at 6 of 11 markers; only two N-block markers reached 10 reads, below the minimum of three, so it was left unassigned even though its typed markers (m.489C, m.8701G, m.10873C, m.14783C, m.15043A, m.16223T) are all consistent with M.

Samples 402 and 431 carry m.10398G, a recurrent position, but four other N-block markers place them inside N.

### **Supplementary Table S12. Allele calls at the eleven diagnostic macrohaplogroup markers**

| **Sample** | **m.8701 (N)** | **m.9540 (N)** | **m.10398 (N)** | **m.10873 (N)** | **m.15301 (N)** | **m.489 (M)** | **m.10400 (M)** | **m.14783 (M)** | **m.15043 (M)** | **m.12705 (R)** | **m.16223 (R)** | **Lowest major-allele fraction** |
| --- | --- | --- | --- | --- | --- | --- | --- | --- | --- | --- | --- | --- |
| 387 | A (284) | T (65) | A (117) | T (219) | G (155) | T (41) | C (138) | T (239) | G (108) | C (125) | C (233) | 1.000 |
| 388 | A (334) | T (126) | A (191) | T (319) | G (258) | T (67) | C (206) | T (357) | G (252) | C (236) | C (720) | 0.997 |
| 389 | G (966) | C (389) | G (576) | C (894) | A (722) | C (248) | T (563) | C (1237) | A (522) | T (616) | T (1908) | 0.997 |
| 390 | G (305) | C (149) | G (202) | C (312) | A (251) | C (81) | T (202) | C (474) | A (245) | T (236) | T (705) | 0.997 |
| 391 | A (282) | T (85) | A (147) | T (293) | G (196) | T (34) | C (160) | T (317) | G (159) | C (181) | C (502) | 0.998 |
| 392 | G (281) | C (111) | G (161) | C (267) | A (252) | C (75) | T (156) | C (368) | A (190) | T (194) | T (901) | 0.993 |
| 393 | A (1484) | T (646) | A (914) | T (1379) | G (1182) | T (220) | C (945) | T (1666) | G (1070) | C (1085) | C (3192) | 0.996 |
| 394 | G (810) | C (362) | G (549) | C (795) | A (654) | C (256) | T (542) | C (1115) | A (628) | T (682) | T (1401) | 0.995 |
| 395 | A (963) | T (282) | A (435) | T (877) | G (638) | T (186) | C (461) | T (951) | G (576) | C (517) | C (1146) | 0.995 |
| 396 | A (237) | T (85) | A (146) | T (198) | G (180) | T (27) | C (148) | T (249) | G (178) | C (160) | C (261) | 0.992 |
| 397 | A (242) | T (97) | A (141) | T (209) | G (181) | T (49) | C (151) | T (289) | G (166) | C (162) | C (300) | 0.990 |
| 398 | G (263) | C (120) | G (161) | C (229) | A (225) | C (76) | T (159) | C (365) | A (188) | T (219) | T (714) | 0.996 |
| 399 | G (777) | C (481) | G (577) | C (824) | A (783) | C (239) | T (566) | C (1257) | A (699) | T (730) | C (1885) | 0.997 |
| 400 | G (131) | C (83) | G (107) | C (134) | A (144) | C (47) | T (105) | C (264) | A (126) | T (152) | T (363) | 0.993 |
| 401 | A (160) | T (74) | A (109) | T (132) | G (106) | T (29) | C (115) | T (174) | G (121) | C (113) | C (81) | 0.991 |
| 402 | A (1390) | T (612) | G (936) | T (1308) | G (1285) | T (669) | C (922) | T (1735) | G (1348) | C (1144) | C (2629) | 0.999 |
| 403 | A (33) | nc (5) | A (11) | T (41) | G (18) | nc (7) | nc (7) | T (29) | nc (8) | C (18) | C (16) | 0.966 |
| 404 | A (148) | T (38) | A (58) | T (128) | G (107) | T (19) | C (61) | T (142) | G (61) | C (56) | C (332) | 0.983 |
| 405 | A (1097) | T (316) | A (487) | T (1050) | G (726) | T (199) | C (516) | T (1164) | G (583) | C (547) | C (1965) | 0.997 |
| 406 | A (350) | T (90) | A (120) | T (306) | G (194) | T (46) | C (136) | T (343) | G (158) | C (147) | C (596) | 0.978 |
| 407 | G (12) | nc (2) | nc (3) | C (13) | nc (9) | C (13) | nc (3) | C (14) | A (12) | nc (8) | T (46) | 1.000 |
| 408 | A (792) | T (181) | A (269) | T (575) | G (507) | T (171) | C (284) | T (719) | G (324) | C (308) | C (1241) | 0.998 |
| 409 | A (143) | T (50) | A (60) | T (163) | G (118) | T (32) | C (66) | T (160) | G (86) | C (60) | C (854) | 1.000 |
| 410 | G (770) | C (349) | G (587) | C (715) | A (617) | C (184) | T (584) | C (1047) | A (627) | T (602) | T (1053) | 0.997 |
| 411 | G (225) | C (95) | G (105) | C (181) | A (166) | C (69) | T (102) | C (332) | A (137) | T (160) | T (519) | 0.996 |
| 412 | G (171) | C (65) | G (100) | C (132) | A (123) | C (46) | T (92) | C (233) | A (135) | T (97) | T (311) | 0.987 |
| 413 | G (240) | C (110) | G (176) | C (255) | A (186) | C (54) | T (165) | C (310) | A (172) | T (199) | T (506) | 0.991 |
| 414 | G (1686) | C (774) | G (965) | C (1634) | A (1110) | C (423) | T (924) | C (2488) | A (1056) | T (1049) | T (2178) | 0.998 |
| 415 | G (457) | C (292) | G (433) | C (514) | A (490) | C (93) | T (419) | C (746) | A (379) | T (472) | T (1205) | 0.996 |
| 416 | A (543) | T (236) | A (383) | T (519) | G (420) | T (162) | C (407) | T (568) | G (380) | C (395) | C (167) | 0.998 |
| 417 | A (768) | T (247) | A (431) | T (652) | G (427) | T (85) | C (477) | T (709) | G (462) | T (420) | T (981) | 0.998 |
| 418 | A (2688) | T (1161) | A (1628) | T (2338) | G (1707) | T (416) | C (1771) | T (2713) | G (1843) | C (1655) | C (2871) | 0.998 |
| 419 | G (83) | C (26) | G (40) | C (81) | A (62) | C (20) | T (36) | C (103) | A (40) | T (42) | T (150) | 0.976 |
| 420 | A (287) | T (75) | A (82) | T (306) | G (193) | T (35) | C (88) | T (317) | G (109) | C (112) | C (747) | 1.000 |
| 421 | G (355) | C (140) | G (200) | C (397) | A (235) | C (115) | T (204) | C (535) | A (230) | T (241) | T (1014) | 0.991 |
| 422 | A (208) | T (76) | A (135) | T (193) | G (150) | T (21) | C (147) | T (223) | G (143) | C (145) | C (251) | 1.000 |
| 423 | A (293) | T (96) | A (199) | T (276) | G (209) | T (58) | C (213) | T (301) | G (195) | C (209) | C (388) | 0.995 |
| 424 | G (759) | C (225) | G (342) | C (654) | A (421) | C (214) | T (325) | C (1092) | A (348) | T (333) | T (1093) | 0.998 |
| 425 | G (4503) | C (2030) | G (2463) | C (4181) | A (2819) | C (1152) | T (2388) | C (6459) | A (2442) | T (2628) | T (7073) | 0.998 |
| 426 | G (97) | C (55) | G (77) | C (89) | A (101) | C (31) | T (75) | C (121) | A (79) | T (87) | T (194) | 1.000 |
| 427 | A (105) | T (20) | A (43) | T (85) | G (77) | T (15) | C (50) | T (83) | G (49) | C (34) | C (127) | 0.987 |
| 428 | G (886) | C (326) | G (556) | C (861) | A (557) | C (214) | T (522) | C (1255) | A (461) | T (546) | T (1741) | 0.995 |
| 429 | A (114) | T (28) | A (44) | T (83) | G (45) | T (12) | C (44) | T (97) | G (49) | C (45) | C (127) | 1.000 |
| 431 | A (1255) | T (324) | G (561) | T (1150) | G (716) | T (658) | C (561) | T (1249) | G (601) | C (659) | C (1075) | 0.997 |
| 432 | G (385) | C (136) | G (181) | C (398) | A (271) | C (84) | T (178) | C (537) | A (197) | T (243) | T (550) | 0.996 |
| 433 | G (279) | C (84) | G (153) | C (258) | A (194) | C (78) | T (151) | C (307) | A (151) | T (200) | T (161) | 1.000 |
| 434 | G (748) | C (437) | G (632) | C (761) | A (666) | C (185) | T (616) | C (1089) | A (674) | T (703) | T (1294) | 0.995 |

Each cell gives the major allele and the number of reads passing filters (in parentheses); nc, not typed (<10 reads).

Letters in column headers indicate the split the marker informs.

Base quality ≥20 and mapping quality ≥30 were required. 508 of 517 markers were typed across 47 samples; no marker was classified as mixed, and the lowest major-allele fraction among typed markers was 0.966, so no sample showed the intermediate fractions expected of a mixture.

### **Supplementary Table S13. Software versions and analysis parameters**

| **Item** | **Tool or parameter** | **Version or value** |
| --- | --- | --- |
| Workflow | Snakemake | 8.14.0 |
| Container | Docker image mtdna-pipeline | 1.5.0 |
| Read trimming | fastp | 0.23.4 |
| Alignment | BWA-MEM | 0.7.18 |
| Alignment processing, pileup | SAMtools | 1.20 |
| Taxonomic classification | Kraken2 (Standard-8, 2024-05-26) | 2.1.3 |
| Scripting | Python | 3.11.9 |
| Alignment access | pysam | 0.22.1 |
| Numerics | NumPy | 1.26.4 |
| Statistics | SciPy | 1.13.1 |
| Figures | Python / matplotlib | 3.13.15 / 3.10.0 |
| Mitochondrial reference | rCRS, NC_012920.1 | 16,569 bp |
| Human reference | GRCh38 (UCSC hg38, no ALT contigs) | — |
| Population annotation | gnomAD v3.1 chrM reduced annotations | md5 45a91b22… |
| Minimum mapping quality | min_mapq | 30 |
| Minimum base quality | min_baseq | 20 |
| Minimum depth to call | min_depth_call | 50 |
| Minimum alternate reads | min_alt_reads | 4 |
| Minimum VAF | vaf_floor | 0.01 |
| Homoplasmy threshold | vaf_homoplasmy | 0.95 |
| Significance level | sig_level | 0.05 |
| Strand-bias rejection | Fisher exact P | ≤ 0.01 |
| Error-model background | VAF below | 0.005 |
| Error model | error_model_pooled | true (per substitution) |
| Beta-binomial concentration bound | α + β | 10⁶ |
| Minimum observations per class | — | 500 |
| Panel of normals, minimum other samples | min_loo_samples | 3 |
| Masked regions | mask_regions | m.302–315; m.16184–16193 |
| Kraken2 exclusion | kraken_exclude | Bacteria, Archaea, Viruses |
| Known-polymorphism AF | common_af | 0.001 |
| Empirical detection limit | applied post hoc | 5% VAF |

The complete configuration file and container definition are archived at <https://doi.org/10.5281/zenodo.23034211> .
